# Functional Analysis of G6PD Variants Associated With Low G6PD Activity in the All of Us Research Program

**DOI:** 10.1101/2024.04.12.24305393

**Authors:** Nicholas R. Powell, Renee C. Geck, Dongbing Lai, Tyler Shugg, Todd C. Skaar, Maitreya Dunham

## Abstract

Glucose-6-phosphate dehydrogenase (G6PD) protects red blood cells against oxidative damage through regeneration of NADPH. Individuals with *G6PD* polymorphisms (variants) that produce an impaired G6PD enzyme are usually asymptomatic, but at risk of hemolytic anemia from oxidative stressors, including certain drugs and foods. Prevention of G6PD deficiency-related hemolytic anemia is achievable through *G6PD* genetic testing or whole-genome sequencing (WGS) to identify affected individuals who should avoid hemolytic triggers. However, accurately predicting the clinical consequence of *G6PD* variants is limited by over 800 *G6PD* variants which remain of uncertain significance. There also remains significant variability in which deficiency-causing variants are included in pharmacogenomic testing arrays across institutions: many panels only include c.202G>A, even though dozens of other variants can also cause G6PD deficiency. Here, we seek to improve *G6PD* genotype interpretation using data available in the All of Us Research Program and using a yeast functional assay. We confirm that *G6PD* coding variants are the main contributor to decreased G6PD activity, and that 13% of individuals in the All of Us data with deficiency-causing variants would be missed if only the c.202G>A variant were tested for. We expand clinical interpretation for *G6PD* variants of uncertain significance; reporting that c.595A>G, known as G6PD Dagua or G6PD Açores, and the newly identified variant c.430C>G, reduce activity sufficiently to lead to G6PD deficiency. We also provide evidence that five missense variants of uncertain significance are unlikely to lead to G6PD deficiency, since they were seen in hemi- or homozygous individuals without a reduction in G6PD activity. We also applied the new WHO guidelines and were able to classify two synonymous variants as WHO class C. We anticipate these results will improve the accuracy, and prompt increased use, of *G6PD* genetic tests through a more complete clinical interpretation of *G6PD* variants. As the All of Us data increases from 245,000 to 1 million participants, and additional functional assays are carried out, we expect this research to serve as a template to enable complete characterization of G6PD deficiency genotypes. With an increased number of interpreted variants, genetic testing of *G6PD* will be more informative for preemptively identifying individuals at risk for drug- or food-induced hemolytic anemia.

## INTRODUCTION

The glucose-6-phosphate dehydrogenase (G6PD) enzyme is expressed in most tissues of the human body^1,2^ but is most notably known for its role in the protection of red blood cells against oxidative damage through regeneration of NADPH and maintenance of reduced glutathione responsible for neutralizing reactive oxygen species.^2^ G6PD deficiency is a common inherited enzymopathy in which a G6PD enzyme with reduced function predisposes individuals to hemolytic anemia upon oxidative stress from drugs, foods, or other insults.^3^ The prevalence of G6PD deficiency ranges from 0 to over 20 percent depending on the geographic region^4^ and is caused by genetic variants in the *G6PD* gene that decrease the function of the G6PD enzyme.^3^ *G6PD* is located on the X chromosome, so individuals who are hemi- or homozygous for variant alleles that produce G6PD with reduced function are at higher risk of acute hemolytic anemia after exposure to oxidative stressors.^5^ Heterozygous individuals can also be at risk since total G6PD activity can vary over time due to variability in X-inactivation,^3^ and identification of carriers is key to determine risk for hemizygous fetuses during pregnancy.^5^ Many G6PD-deficient individuals live without major negative health consequences until exposure to oxidative stressors, but when encountered, these can result in serious hemolytic anemia requiring urgent care.^5^ Thousands of deaths worldwide can be attributed to G6PD deficiency every year,^6,7^ highlighting the importance of prevention and medical management of G6PD deficiency-related hemolytic anemia. Prevention of G6PD deficiency-related hemolytic anemia can be readily achieved through genetic testing or G6PD activity assays and subsequent avoidance of offending agents (e.g. fava beans, dapsone, rasburicase, primaquine, etc.).^3^

While G6PD activity assays are the gold-standard for diagnosing G6PD deficiency, they cannot be used in the midst of a hemolytic crisis since the red blood cells with lowest G6PD activity will have already been lost to hemolysis, so it is recommended to wait three months before re-testing to make a diagnosis of G6PD deficiency.^5,8^ In contrast, genotyping or sequencing of the *G6PD* gene can be performed at any time, ideally before an adverse reaction is experienced, to provide a clinically valid estimation of G6PD deficiency.^3^ As the number of individuals with available whole-genome sequencing (WGS) or pharmacogenomic genotyping results increases, genetics will become a more accessible and less invasive method for the identification of individuals with G6PD deficiency. This is exemplified by the Medicine and Your DNA opt-in return-of-results to All of Us participants that is underway by the All of Us Research Program.^9^ Returning genetic test results with clinical interpretations to patients and medical providers relies on variant interpretations that are based on sound evidence; however, of over 1300 identified variants in G6PD, more than 800 have too little evidence to use for diagnosis of G6PD deficiency.^10^ This is especially true for rare variants, so this gap can be addressed in part through the analysis of large cohorts of diverse individuals with WGS and linked medical records, such as that available from the All of Us Research Program.^11^ This approach becomes especially powerful when combined with *in vitro* functional assays.

There also remains significant variability in which G6PD deficiency-causing variants are included in pharmacogenomic testing arrays across institutions, many only including c.202G>A (with or without the c.376A>G variant, which together are the most common G6PD-deficiency allele, G6PD A-).^12–14^ However, dozens of other variants can also cause G6PD deficiency and thus would lead to false negative results for many individuals if using one of these minimal arrays.^3^ This potential deficit can be assessed using data available in the All of Us Research Program to identify how many individuals with G6PD deficiency-causing variants would not be identified by arrays containing a limited set of *G6PD* variants.

Interpretation of *G6PD* variants, based on the degree to which they cause G6PD deficiency, is important to advancing the clinical utility of *G6PD* genetic testing, and systems designed to classify variants have been evolving. In 1989 a World Health Organization (WHO) panel established a classification system for *G6PD* alleles, where Class I variants contribute to chronic anemia, Class II and III lead to G6PD deficiency with risk of acute hemolytic anemia, and Class IV have normal activity.^15^ They revised this system in 2022 such that Class A contribute to chronic anemia, Class B reduce activity below 45% and increase risk of acute hemolytic anemia, and Class C have over 60% normal activity and are presumed normal-like; any intermediate variants or variants without activity measurements in three unrelated hemizygous individuals are defined as uncertain, in Class U.^16^ The American College of Medical Genetics (ACMG) variant interpretation framework is another system that is used for *G6PD* variant interpretation. The ACMG framework scores pieces of evidence to determine if there is sufficient support that a variant is pathogenic and contributes to a specific condition, or is benign and does not contribute.^17^ A ClinGen panel of experts is working to adapt the ACMG framework specifically for G6PD.^18^ These efforts by the WHO and ClinGen rely on clinicians, researchers, and genetic testing companies to generate additional evidence that enables classification and interpretation of more *G6PD* variants.

In this study, we aimed to contribute to the global efforts to improve *G6PD* genotype interpretation by describing the association between *G6PD* variants identified in the All of Us Research Program and G6PD deficiency phenotypes, including the epidemiology associated with each *G6PD* coding sequence (CDS) variant. We performed functional assays to confirm the activity of any unclassified variants with measured G6PD activity in All of Us individuals. Our results increase the number of variants that can be classified into non-uncertain categories and demonstrate that expanding the number of *G6PD* variants included on pharmacogene testing arrays, or utilizing whole-genome sequencing, is necessary to identify more G6PD deficient individuals.

## METHODS

### All of Us Research Project Genotype Determination

The All of Us version 7.1 release contains short read WGS data for 245,394 individuals, organized into several data formats including a convenient subset of exonic regions with variant calls organized into Plink files. These exonic regions include UTRs and a padding of 15 bases into surrounding intron regions to catch splice-site relevant variants.

Information about how the All of Us organization calls variants from the sequencing data can be found in the All of Us documentation.^19^ We first downloaded the X chromosome Plink files into the cloud computing environment. We then addressed sex assignment since it underlies the assignment of homozygous vs. hemizygous labels to *G6PD* variant alleles and to set heterozygous haploid calls as missing. We extracted the survey response results for “sex at birth” which included 94,756 male, 145,563 female, and 5,075 responses other than male or female. We extracted the non-pseudoautosomal regions from the hg38 base pair coordinates 2,781,479 to 155,701,383 resulting in 941,048 variants. Using the Plink “—impute-sex” command, we retrieved the X chromosome homozygosity rates (F coefficient = observed homozygosity/expected homozygosity)^20^ for each individual, as those with high rates of homozygosity (reference allele or variant) are the ones with only one X chromosome. F values between 0.2 and 0.8 represent potentially ambiguous calls, of which there were 94 individuals. To assign sex to these individuals, we iteratively tested every F value threshold, at increments of 0.01, between 0.2 and 0.8 to find a single threshold that would result in the greatest concordance with the survey-based “sex at birth”, finding 0.77 was the best threshold. Plotted on a density plot (**Supplemental Figure S1**), 0.77 was visually the best separation between the two distributions of F values, thus, individuals below 0.77 were assigned as having more than one X chromosome (148,489 individuals) and individuals above or equal to 0.77 were assigned as having one X chromosome (96,905 individuals). This method resulted in >99.99% concordance with the survey responses that were either male or female. There were 12 discordant results which were all from male survey responses with lower than 0.77 homozygosity rates. There remained only 1 discordant result when compared to the All of Us Dragen ploidy determinations as described in the QC metrics. Since our method of sex determination using Plink’s F coefficients resulted in sex assignments for all individuals (whereas Dragen leaves many ambiguous) and had >99.99% concordance with Dragen and survey-based sex at birth, we used the genetically determined homozygosity rates described here to assign sex to the Plink files. As an additional quality control measure, we inspected the heterozygous haploid calls and found every individual assigned as having one X chromosome (“male”) had at least one heterozygous variant genotype, indicating that there is some inaccuracy in the genotypes produced from the sequencing data which is described in the QC documentation provided by All of Us.^19^ The most heterozygous haploid calls for any individual was 246 out of a total of 941,048 variants in the data, but this individual had 572 homozygous minor alleles. Since it is typically expected to observe more heterozygous alleles than homozygous minor alleles in a diploid individual (and we observed the inverse of this) we are confident that all individuals assigned as having one X chromosome are accurate. Therefore, we considered heterozygous haploid genotypes as missing for the remaining analyses.

From these files, a parallel set of Plink files was created by filtering to the Ensembl-defined canonical *G6PD* transcript coding sequence (CDS, provided in **Supplemental Table S1**) and filtering to variants that were present in at least 1 allele (Plink argument “--mac 1”). There were 27 heterozygous haploid calls across 27 individuals and 7 variants. These were set to missing.

Intronic variants and variants surrounding the *G6PD* gene were extracted from the All of Us VariantDataset using Hail version 0.2.126 and converted to Plink files. The same sex assignments were applied to this data.

### All of Us Research Project Phenotype Determination

Four phenotypes were curated: (1) G6PD activity assay, (2) diagnosis of any hemolytic anemia, (3) diagnosis of G6PD deficiency with or without anemia, and (4) diagnosis of drug induced hemolytic anemia with or without an autoimmune component.

Figure 1 shows the steps taken to curate the activity assay results from nine distinct laboratory value concept names containing “G6PD” or “Glucose-6-Phosphate dehydrogenase”. The G6PD genetic testing field was removed since the data did not indicate the resulting genotypes, only that the test was performed. Since G6PD activity assays are not standardized across institutions, values reported in the All of Us data must be scaled to reference ranges reported by the testing lab. Unfortunately, more than half the data did not contain reference ranges or reliable test units (e.g. units per gram hemoglobin), requiring inferences to be made regarding the “normal activity” scalar required to derive the percent-activity-of-normal results. Extensive data exploration revealed that some of the normal ranges were in the 200’s, but with the majority below 30. The concept codes (provided in **Supplemental Table S2**) and reported assay units do not adequately enable grouping activity measurements into groups corresponding to the type of G6PD assay the value was derived from (“like-for-like” groups), requiring a stepwise curation strategy described in the text boxes of Figure 1. This involved finding criteria that enabled grouping measurements into like-for-like groups, and inferring the most appropriate reference range for that group. For example, values that contained decimal points in the reported value and had corresponding reference ranges, all had reference ranges below 21. This enabled the inference that values with decimal points *without* reference ranges could be compared to the median of the reference ranges that exist in the rest of the values with decimal points and with reference ranges. Individuals for whom values were derived based on inferred reference ranges are flagged in the results as “inferred” as there is the potential for some values to be biased low or high if the true reference range differed from the inferred. For example, an activity value of 4 would be assigned 50% of normal if compared to a normal range of 6-10, and 25% if compared to a normal range of 13-19.

**Figure 1:**
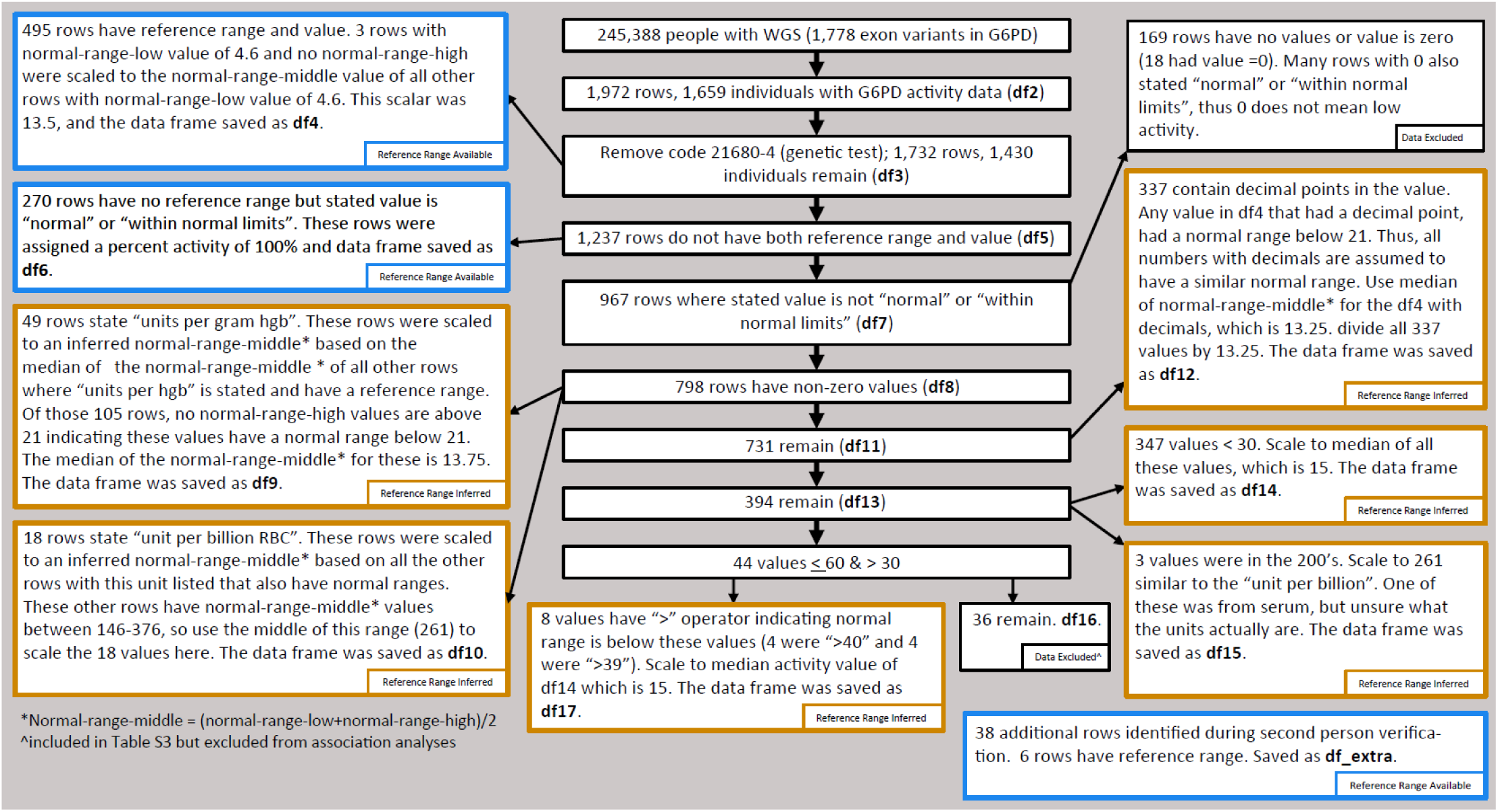
Phenotype curation method for G6PD activity data in the All of Us v7.1 release. “Rows” refers to a line of data where an activity result is reported. There can be multiple rows of data for an individual if they had multiple G6PD activity assays recorded in their medical record. Although there are 245,394 individuals with short read WGS, six individuals do not have Curated Data Repository (CDR) data, resulting in 245,388 individuals for phenotype curation.

Each separate assumption is detailed in Figure 1 and each box corresponds to a data frame that was inspected for bias. The distributions for each of these groups is provided in **Supplemental Figure S2**. Two groups were excluded based on biases or inferences that were unresolvable. This includes the values that were zero or missing and did not state “normal” or “within normal limits”. The other group consists of values between 30 and 60, without “>” operators, and these were deemed unresolvable because they could have been low values with a reference range in the hundreds or high values with a reference range below 30. However, this group was included for the purpose of **Supplemental Table S3**, but excluded for all other analyses. To derive percent-activity-of-normal for each like-for-like dataset, the activity values were divided by the middle (mean of the low and high reference) of the available or inferred reference range and log2 transformed. The data frames were combined and the lowest value for each individual was used. Visualization of the data from inferred reference ranges distributes similarly as the data with available reference ranges, with only slightly more variability (shown in **Supplemental Figure S2**, and further discussed in the results section), providing confidence to the inferred reference range data.

Phenotypes represented by diagnosed conditions were extracted using standard concept names which are related to international classification of disease (ICD) diagnosis. Twenty-five (25) standard concept names exist under the parent concept “hemolytic anemia” in the All of Us Research Program health data records. If an individual had any instance of a diagnosis of any type of hemolytic anemia, they were considered cases for the corresponding analysis. These diagnosis names are provided in **Supplemental Table S4** and include drug-induced hemolytic anemia. G6PD diagnosis was coded in the same way, for two concepts: “G6PD deficiency with anemia” or “G6PD deficiency without anemia” (**Supplemental Table S2**).

After phenotypes were assigned, the corresponding Plink files were created to include individuals (1) with WGS, (2) with activity data, (3) with a diagnosis of hemolytic anemia, (4) with a diagnosis of G6PD deficiency, and (5) with a diagnosis of drug induced hemolytic anemia. These Plink files were converted to .raw files (rows are individual identifiers and columns are variants) and defined the alleles-to-count (i.e. the variant alleles) as the minor allele from the total WGS data. The number of heterozygotes, hemizygotes, and homozygotes in each group was counted under an additive model. Hemizygosity or homozygosity for each variant in these files were assigned the number 2, while heterozygosity was assigned the number 1, and the reference genotypes (defined by the allele with the highest allele frequency) were assigned 0. For calculations of allele frequencies and allele counts, hemizygous variants were counted as 1 variant allele in a total of 1 X chromosome. **Supplemental Table S3** specifically lists each Class I/II/III or Class A/B variant that was found in individuals with G6PD activity assay results and the other G6PD deficiency-related phenotypes. There were other Class I/II/III or Class A/B variants that were present only in individuals with WGS data yet without a G6PD phenotype, and therefore were grouped into a single line “Other Class I/II/III or Class A/B” in **Supplemental Table S3**.

### Haplotype Estimation

We used Aldy version 4.4^21^ and the gnomAD variant co-occurrence calculator^22^ to determine that the c.542A>T and c.563C>T most commonly exist on independent haplotypes from the c.202G>A; thus, individuals with both c.202G>A and c.542A>T or c.563C>T are likely compound heterozygotes. For estimating the likely haplotypes and diplotypes for other individuals, the c.202G>A and c.376A>G variants were assumed to be on the same haplotype when both genotypes were present, the c.376A>G and c.968T>C assumed to be on the same haplotype, and the c.1048G>C assumed to be on independent haplotypes from c.542A>T or c.376A>G.

### Variant Classification According to WHO Criteria

To define variants by WHO class and annotate variant consequences from the existing knowledge base, we used supplementary tables originally published in Geck et. al^16^ and maintained at https://github.com/reneegeck/G6PDcat, accessed October 27, 2023. In these tables the c.1311C>T (p.Tyr437=) was determined to fit the Class B category based on a few cited reports of decreased activity, but since c.1311C>T is found in over 10% of individuals in gnomAD and reported to be benign (ClinVar Variation ID 470162), we excluded this variant from the Class B category for all further analysis. Further analysis described in the results section of this manuscript provides evidence to support the non deficiency-causing nature of this variant, which we submitted to update the aforementioned tables. For variants not included in this annotation file, we used the UCSC Variant Annotation Integrator^23^ to retrieve variant consequence and other details.

### All of Us Research Project Associations Between Genotype and G6PD Diagnosis

To determine if novel variants associate with individuals with a deficiency diagnosis who didn’t have a known genetic cause of G6PD deficiency, we performed a Fisher exact test comparing the allele frequencies of all *G6PD* variants found in 19 individuals with a deficiency diagnosis who didn’t have WHO Class I/II/III or A/B variants (excluding c.1311C>T) versus 1097 individuals who had normal (> 60%) G6PD activity and no WHO class I/II/III or class A/B variant (excluding c.1311C>T). Bonferroni correction was used to correct for multiple testing.

### All of Us Research Project Associations Between Genotype and G6PD Activity

We performed a chromosome X exonic association study (XWAS) among 1,276 unique individuals with interpretable activity values as described in Figure 1. This analysis tested 16,434 variants restricted to variants with over 90% genotyping rate and present in 3 or more alleles. A similar analysis was conducted for all variants in the short-read WGS data for the region of Chromosome X +/-1 million base pairs (bp) from the *G6PD* region (chrX:153527010-155552570, hg38), resulting in 11,682 tested variants, restricted to variants with over 90% genotyping rate and present in 3 or more alleles. For restricting the analysis to 3 or more alleles, we confirmed that hemizygous males were counted as haploid even though the genotype is counted as 2 for the linear regression. The association studies were conducted under an additive model assuming heterozygotes can have slightly decreased G6PD activity. The Plink (version 2.0) command “— assoc” was used to perform the linear regressions between the log2 transformed activity values and each genotype. The “—assoc” command is equivalent to specifying that haploid genotypes be set to 0 or 2, and to not include sex as a covariate. Covariates were intentionally not included in the analysis because there was no intent to control for environmental variables given the direct effect of *G6PD* variants on G6PD activity, and since G6PD activity is already normalized to red blood cell counts by the testing lab. Leaving out covariates also simplifies the interpretation of the results, as resulting statistical estimates are not adjusted for other variables. The package qqman (version 0.1.9)^24^ was used to visualize QQ plots and plot the Manhattan plots.

The methodology we employed to correct for the effect of known loss-of-function *G6PD* variants was to exclude individuals who have a clear genetic basis for lower G6PD activity. Therefore, corrected association studies were completed after removing individuals hemizygous, homozygous, or compound heterozygous for WHO class I/II/III or A/B variants (not including c.1311C>T). Heterozygotes were not removed to preserve sample size and since only a small effect size is expected in these individuals. However, the c.542A>T and c.202G>A alleles were present in one suspected compound heterozygote (who would therefore be expected to have G6PD deficiency) whose WGS data was not able to be phased due to being ∼2000 bp apart (no long-read sequencing data was available for this individual). GnomAD allele combinations indicate that these variants most likely exist on different haplotypes, and this agrees with the diplotype called for this individual by Aldy^21^ version 4.4 (A-/ Santa Maria), which uses population-based phasing to infer haplotypes. Due to the evidence towards compound heterozygosity, we removed this compound heterozygote along with individuals homozygous or hemizygous for the WHO class I/II/III or A/B variants. This resulted in 1,247 individuals with 16,302 variants for the XWAS and 11,626 variants for the +/-1 million bp *G6PD* region association study.

To control for the increased probability of false positives with multiple testing, we employed permutation testing to determine the most appropriate significance threshold at an alpha of 0.05. This was done by randomly scrambling the phenotype column (G6PD activity) using the base R command “sample” and repeating the analysis 1000 times. The lowest p-value from each permutation was recorded and the value defining the fifth percentile of this resulting distribution was set as the 5% false discovery threshold. This is the most robust and empirical method available to define this threshold.^25–27^

To determine what the G6PD activity association study was powered to detect, we employed a custom-built code to simulate the exact conditions of our study for genetic variants with a spectrum of effect sizes in a population representative of the actual All of Us cohort under analysis. We built a population 100 times larger than our sample size (synthetic population = 124,700) with each variant present in the same zygosity proportions as our actual cohort. We pruned the variants down to just the ones with unique allele frequencies and unique zygosity proportions so as to not unnecessarily duplicate iterations of the loop. Instances of undetermined genotypes “NA” from the actual All of Us population were assumed to be homozygous reference allele (the one with the highest allele frequency) for the purpose of reproducing the genotype distributions. For each variant, using the Faux package (version 1.2.1; command “rnorm_pre”),^28^ a phenotype was synthesized which matched the actual All of Us distribution of G6PD activity values (mean = -0.059, standard deviation = 0.443), and forced to have the defined correlation with the variant in the population. The synthesized phenotypes were set to have correlations of -0.1, -0.2, -0.22, -0.24, -0.27, -0.3, -0.4, -0.5, - 0.92, and -0.99 with each variant in the synthetic population. For each variant, for each correlation level, we sampled 1,247 individuals from the synthetic population 500 times, conducting linear regression each time, and recording the resulting p-value of each iteration. The fraction of p-values that were higher than (not passing) the significance threshold of 7.7×10^-17^ (determined by permutation testing for the corrected G6PD-WAS) was the estimate of the type II error (false negative rate), and power was determined as 1 minus the false negative rate.

### All of Us Research Project G6PD Activity by Haplotype Analysis

We analyzed the All of Us activity data for 1,255 individuals with interpretable activity values (see Figure 1) and 100% non-missing genotyping calls for *G6PD* CDS missense variants. Less than 2% of the data (21 individuals) were removed based on missing *G6PD* missense genotypes, all of these being genetically inferred males (one X chromosome) with normal activity values and no diagnosis of G6PD deficiency. We do not suspect these individuals were misassigned as X chromosome haploids (males) given the quality control measures described earlier. However, we still chose to exclude these individuals to achieve the highest accuracy possible within this analysis. Every unique combination of *G6PD* CDS missense variants were determined and used to categorize individuals into G6PD groups with the same genotype combination. These were notated by zygosity, CDS position, nucleotide change, and underscores (e.g. “het202G>A_het376A>G”).

We followed the same procedure to analyze the synonymous variant genotype combinations for 1,262 individuals with interpretable activity values and 100% non-missing genotyping calls for *G6PD* CDS synonymous variants.

### Cellular Functional Studies of Variant Consequences to G6PD

*S. cerevisiae* lacking *ZWF1* (*zwf1Δ*), strain YMD4438 (**Supplemental Table S5**), were transformed with a plasmid for expression of a human *G6PD* variant under control of the *S. cerevisiae CYC1* promoter (p416CYC1); each plasmid also contained the *URA3* gene to enable selection by growth in sc-ura media.^29^ For transformations, YMD4438 was grown overnight in YPD (1% yeast extract, 2% peptone, and 2% dextrose). Per transformation, 1mL of overnight culture was pelleted, 100ng of plasmid DNA added, followed by 75µL of transformation buffer (0.2M lithium acetate, 0.1M DTT, 40% PEG3500, 0.5mg/mL carrier DNA). After 1 hour incubation at 42°C, the transformation reaction was plated on sc-ura and incubated 3 days at 30°C. Two transformants were selected per plasmid. For the functional assay, the resultant strains (**Supplemental Table S5**) were grown overnight in sc-ura, and 2µL was inoculated into 198µL of indicated media (control sc-ura; selection sc-ura-met +1mM hydrogen peroxide) in a 96-well plate (Corning CLS3370). Optical density at 600nm was measured every 15 minutes on a BioTek Synergy H1 with orbital shaking at 30°C. Growth curves were plotted in R, and area under the curve was computed using gcplyr.^30^ The selective growth condition was chosen for its ability to differentiate variants associated with G6PD deficiency, with activity ranging from 1-25% of normal,^10^ from the reference variant.

### Variant Effect Prediction Tools

Missense variant pathogenicity computational prediction tools were selected from tools tested for ClinGen recommendations.^31^ Scores for ten of these tools were available from dbNSPF v4.^32,33^ ENST00000393562 was used for all tools for which it was available (BayesDel, CADD, GERP++, PrimateAI, REVEL, MPC, PolyPhen-2 HumVar), and ENST00000393564 was used for the remaining tools (SIFT, FATHMM, VEST4). Scores were converted to interpretations based on ClinGen recommendations.^31^ Scores from FATHMM and CADD were not included in final analysis because they predicted all tested variants to be pathogenic and benign, respectively. If other tools predicted conflicting evidence for benignity or pathogenicity, consensus was taken when more than ⅔ agreed and no more than one tool dissented, and the consensus was used at supporting level of evidence. If multiple tools provided non-conflicting evidence, the lowest level of support was conservatively used.

We also used the dbNSFP v3^34^ accessed through the UCSC Variant Annotation Integrator^23^ to provide SIFT and PolyPhen-2 in silico predictions of deleteriousness of variants not previously annotated by Geck et. al.

### Variant Interpretation Updating

To update the existing knowledge base with findings reported in this study, variant interpretation guidelines were applied according to Richards et al.^17^ We applied these variant interpretation guidelines to any variants of uncertain significance or conflicting interpretations on ClinVar that were found in individuals with G6PD activity measurements. Specifically, measurement of decreased G6PD activity was used with evidence code PP4; activity within the normal range was used for BS3 or if the normal range was inferred was modified to be at supporting level of evidence, denoted as BS3_P. Significantly reduced growth in our functional assay was used for PS3. Variant effect predictions were used for PP3 and BP4 as described above. All other codes were adapted as previously reported;^10^ briefly, PM1 was applied to NADP binding domain from residues 38–44, substrate binding domain from residues 198–206, dimerization domain from residues 380–425, and structural NADP binding site from residues 488–489;^35–38^ PM2 was used with a conservative cutoff for carrier frequency of 0.001 since allele frequency can be below 0.01 depending on the region;^4,39^ BA1 was applied to variants >5% frequency in gnomAD; BS2 was used for reports of nondeficiency without activity measurements provided; BP6 was used for prior reports to ClinVar of benignity. Interpretations will be deposited to ClinVar following peer review, and all variant information is available at https://github.com/reneegeck/G6PDcat.

### Second Person Verification and Data/Code/Strain Availability

The genetic data was independently verified by one of the other authors. The same was done for assignment of phenotypes and inference of reference ranges. Any discrepancies were reviewed and methods adjudicated to result in the methods described in this section.

All code used to generate the All of Us analyses is documented in the All of Us cloud computing platform as step-by-step notebooks in a dedicated workspace. We will share the workspace as read-only with approved users of the All of Us controlled tier data upon request. A documented exception was granted by the All of Us Research Access Board to display data from groups of individuals below sample sizes of 20, including cases where n=1. Code for analysis of *S. cerevisiae* functional assays are available at https://github.com/reneegeck/DunhamLab and strains (**Supplemental Table S5)** are available upon request. Figures were compiled using R and Adobe Illustrator (version 27.1.1).

## RESULTS

### G6PD CDS Variants in the All of Us Population

There were 245,394 individuals with WGS data in the All of Us version 7.1 release (96,904 with one X chromosome, 148,490 with 2 or more), and 38.7% (94,869) had at least one variant in the *G6PD* CDS (for list of variants, see **Supplemental Table S6**). 14.0% of these individuals (13,298; 5.4% of the total cohort) had at least one nonsynonymous variant classified as WHO class I/II/III or A/B, and 927 individuals had at least one unclassified nonsynonymous variant (0.3% of the total cohort).

Narrowing the focus to individuals that would be predicted to have G6PD deficiency, 1.7% (4,077) of all individuals with WGS were homozygous or hemizygous for a WHO class I/II/III or class A/B variant (not including compound heterozygotes). Of these 4,077 individuals, 1.2% (49) had at least one instance of hemolytic anemia diagnosed from any cause, and 0.8% (33) had at least one instance of G6PD deficiency (with or without anemia) diagnosed. This demonstrates that over 99% of individuals homozygous or hemizygous for WHO class I/II/III or A/B variants are not documented (in the available data) with a diagnosis of G6PD deficiency, an enzymopathy they likely have.

**Supplemental Table S3** provides a summary of the variants found in the All of Us data and shows how many heterozygotes and homo/hemizygotes there were with each variant. This table is agnostic of haplotypes and therefore only serves as an overview of how many people had each variant in each of the phenotype groups.

There were 359 *G6PD* CDS variants present in the All of Us v7.1 Research Program participants with WGS (**Supplemental Table S6**). Of these 359 variants, 7 have been previously categorized as WHO class IV or class C and 55 variants have been previously categorized as WHO class I/II/III or class A/B. Among the 4,077 individuals homozygous or hemizygous for these WHO class I/II/III or class A/B *G6PD* variants, 87% were homozygous or hemizygous for the c.202G>A variant, demonstrating that 13% of individuals in this population who likely have G6PD deficiency based on their genetic variants would be missed by only testing for the c.202G>A variant.

296 *G6PD* CDS variants were found in individuals in the All of Us Research Program that were not classified as WHO class I/II/III/IV or A/B/C. Of these, 150 were nonsynonymous variants. These included a frameshift reported to ClinVar as pathogenic (c.1054_1055insG) identified in a heterozygote, and five missense variants reported to ClinVar as likely pathogenic by at least one submitter, though some have conflicting interpretations (c.49C>T, c.193A>G, c.448G>A, c.582C>G, and c.739G>A). We identified 32 nonsynonymous variants (provided in **Supplemental Table S7**) that had not been previously reported to ClinVar or included in curated lists of *G6PD* variants from multiple databases and publications.^10^ Of these nonsynonymous variants without previous annotations, 30 (out of 32) were missense variants; the remaining two were an in-frame deletion identified in a heterozygous individual and a nonsense mutation identified in a heterozygous individual. There were 146 *G6PD* CDS synonymous variants in individuals in the All of Us Research Program that were not classified as WHO class I/II/III/IV or A/B/C.

### Association of G6PD Variants with G6PD Deficiency Diagnosis

To assess the connection between *G6PD* variants and G6PD deficiency, we first analyzed reports in All of Us of a G6PD deficiency diagnosis with or without anemia. There were 82 individuals with a G6PD deficiency diagnosis, yielding a prevalence of 0.03% (82/245,394) in the All of Us Research Program. Among individuals with a diagnosis of G6PD deficiency, only 33 (out of 82) were homozygous or hemizygous for a WHO class I/II/III or class A/B variant. Of the other 49 diagnosed individuals, 30 were heterozygous for WHO class I/II/III or A/B variants (2 of these were likely compound heterozygotes), 12 had no genetic variants in the CDS of *G6PD*, and the remaining 7 individuals only had class IV/C or synonymous variants that are not expected to significantly affect G6PD activity (summarized in **Supplemental Table S3**). In total, there were 11 *G6PD* variants in the CDS of All of Us participants with a G6PD deficiency diagnosis, 8 of which were missense. The missense variant genotype combinations and estimated haplotypes for individuals with a diagnosis of G6PD deficiency with or without anemia and no missing missense genotypes are provided in **Table 1**.

**Table 1:** Missense genotype combinations and estimated haplotypes in individuals with a G6PD deficiency diagnosis with or without anemia. One individual had a missing genotype that was excluded from analysis. Sample sizes are described with information about X chromosome ploidy.

| Estimated Haplotype | Genotype Combination | Number of Individuals by X Chromosome Ploidy |  |
| --- | --- | --- | --- |
|  |  | 1X | 2X |
| A<sup>-(202)</sup> / A<sup>-(202)</sup> | hom202G>A_hom376A>G | 17 | 5 |
| A<sup>-(202)</sup> / B | het202G>A_het376A>G | 0 | 20 |
| B / B | No missense variants | 6 | 10 |
| Mediterranean / Mediterranean | hom563C>T | 7 | 0 |
| A<sup>-(202)</sup> / A<sup>+</sup> | het202G>A_hom376A>G | 0 | 5 |
| A<sup>+</sup> / B | het376A>G | 0 | 2 |
| Mediterranean / B | het563C>T | 0 | 2 |
| Union / Union | hom1360C>T | 1 | 0 |
| Kaiping / Kaiping | hom1388G>A | 1 | 0 |
| Mediterranean / A<sup>-(202)</sup> | het202G>A_het563C>T_het376A>G | 0 | 1 |
| A<sup>-(202)</sup> / Santa Maria | het202G>A_het542A>T_hom376A>G | 0 | 1 |
| Dagua / B | het595A>G | 0 | 1 |
| A<sup>-(968)</sup> / A<sup>+</sup> | het968T>C_hom376A>G | 0 | 1 |
| A<sup>-(968)</sup> / A<sup>-(968)</sup> | hom968T>C_hom376A>G | 1 | 0 |

G6PD deficiency diagnoses in this real-world data are observed in 16 individuals without a clear genetic basis for G6PD deficiency (no missense variants) and 28 individuals heterozygous for missense variants (individuals who are usually unaffected but can have variable activity^3^). All *G6PD* variants (chrX:154531391-154547572) in individuals with a diagnosis of G6PD deficiency are listed in **Supplemental Table S8**. Since there remains the possibility that noncoding *G6PD* variants contributed to these diagnoses in individuals without a clear genetic basis for the disease, we performed a Fisher exact test comparing the allele frequencies of all *G6PD* variants found in 19 individuals with a deficiency diagnosis who didn’t have WHO Class I/II/III or A/B variants versus 1097 individuals who had normal (> 60%) G6PD activity and no WHO class I/II/III or class A/B variant. There were 70 *G6PD* variants tested, the results of which are provided in **Supplemental Table S9**. No variants were statistically significantly enriched in cases after Bonferroni correction. Upon visual inspection of the top results (uncorrected p-values 0.002871 and 0.01785) (**Supplemental Figure S3**), the low number of case counts in individuals with these variants (2 heterozygotes and 2 homo/hemizygotes) do not convey that these findings are strong enough to warrant closer investigation.

### G6PD Activity Assay Phenotype Curation

To enable assessing the connection between *G6PD* variants and G6PD activity, we curated the clinical G6PD activity assay records in the All of Us Research Program data. G6PD activity assay data was available for 1,665 individuals with short-read WGS. To interpret activity values from assays which were conducted at different testing centers, each value needed to be converted to percent of normal activity, and therefore required normal reference values. The normal values, however, differ depending on the specific G6PD testing laboratory and assay methodology used. For 771 activity assays (i.e. a row of data corresponds to an activity assay result), normal ranges were provided to All of Us or the assay was described as “normal” or “within normal limits”, resulting in easily interpretable activity values. For the remaining assay data rows, normal ranges were inferred from similar tests for 762 assays resulting in interpretable activity values, and the remaining 205 assay data rows were excluded from further analysis (Figure 1). This resulted in successful assignment of activity assay values for each individual, as demonstrated by the distribution of log2 transformed percent-of-normal G6PD activity not differing significantly between samples where a normal range was provided (n=660) versus samples where we inferred the normal range (n=616) (p=0.0627 by Welch’s two sample t-test) and the difference in means corresponding to a difference in activity of only 4% between these two distributions. Additionally, the Class I/II/III and A/B variants expected to cause deficiency were observed with low G6PD activity in the inferred results, and class C variants (non-deficiency-causing) were observed with normal G6PD activity in the inferred results, providing further confidence to the results based on inferred normal ranges. **Supplemental Figure S2** shows the distributions of the inferred values versus the ones with normal ranges provided.

### Chromosome X and G6PD -Wide Variant Association Tests with G6PD Activity

We aimed to discover variants that are common or effectual enough to be associated with G6PD activity. To do this, we first confirmed the association between WHO class I/II/III or A/B variants and low G6PD activity by conducting an exonic chromosome-X wide association study (XWAS) using the whole genome sequencing results for 1,276 individuals with interpretable G6PD activity data. Since we do not expect variants far away from the *G6PD* gene to be significantly associated with G6PD activity, this analysis serves to demonstrate that the methodology is effective at differentiating false positives from true signals. This analysis tested 16,434 variants, restricted to variants with over 90% genotyping rate and present in 3 or more alleles, demonstrating that there is a strong association signal with G6PD activity in the *G6PD* gene region (Figure 2A). The strongest association is for the common G6PD deficiency-causing c.202G>A variant, as expected, and another strong association with the known G6PD deficiency-causing c.563C>T variant (both shown in more detail as insets in Figure 2A). No associations above p-value 0.5 are displayed, and variants within the *G6PD* CDS are indicated by green dots in Figure 2.

**Figure 2:**
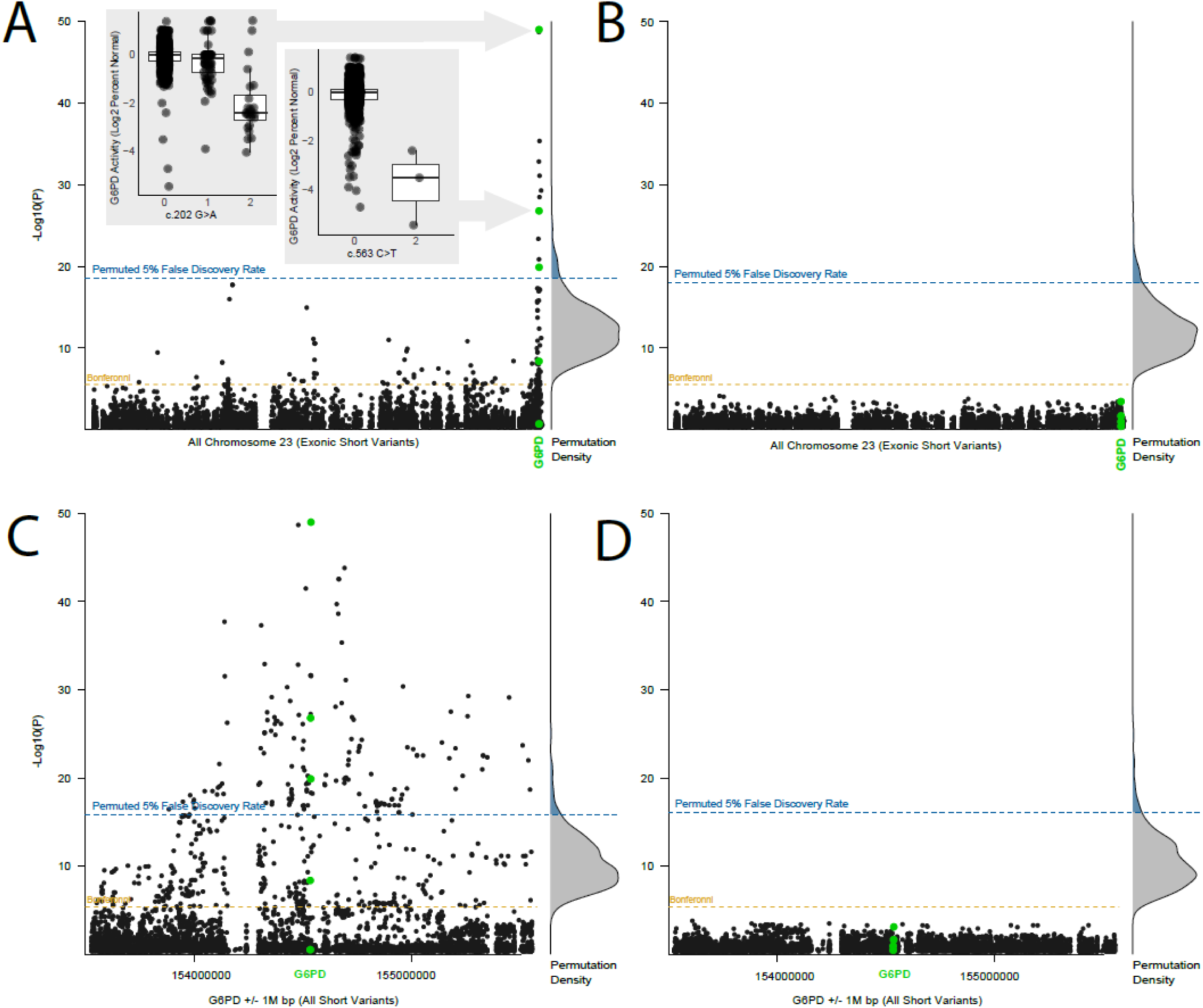
Manhattan plots for G6PD activity-variant associations, filtered to only those with p-values below 0.5. (A) Associations between chromosome X variants in exons (including UTRs and 15 bases into introns) detected by short read WGS and G6PD activity in 1,276 individuals, and (B) The same analysis as A but for 1,247 individuals that are not homozygous, hemizygous, or compound heterozygous for WHO class I/II/III or A/B variants. (C) Associations between variants detected by short read WGS +/-1M bases from the *G6PD* region and G6PD activity in 1,276 individuals, and (D) the same analysis as C but for 1,247 individuals that are not homozygous, hemizygous, or compound heterozygous for WHO class I/II/III or A/B variants. All panels: The gold dotted line is the Bonferroni significance threshold (alpha = 0.05/number of tested variants), and green dots are highlighted variants within the *G6PD* CDS region. 1000 permutations of randomly scrambled phenotype data was used to visualize the p-value distribution of top hits from each random permutation (hits that occur by random chance), shown as density distributions to the right of each Manhattan plot. The 5th percentile in p-values from these permutations empirically quantifies the threshold at which association results have less than 5% chance of being false positives (blue dotted line).

Since many variants passed the Bonferroni significance threshold (above the gold line in Figure 2A), we also used permutation analysis to identify the actual threshold at which false positives would occur at a rate of 5% (see Methods). Based on the more stringent permuted 5% significance threshold, only variants near the *G6PD* gene boundary (*G6PD* locus) remained statistically significant, confirming the effectiveness of permutation testing. We then repeated the analysis after removing individuals homozygous, hemizygous, or compound heterozygous for the WHO class I/II/III or A/B variants. The results of this corrected association test (Figure 2B) demonstrate that many variants that were Bonferroni-significant throughout the X chromosome in Figure 2A disappear, further demonstrating the effectiveness of the employed methodologies to discern true from false positives and demonstrating no new exonic variants in *G6PD* correlate significantly with G6PD activity.

To determine if variants in *G6PD* introns, UTRs, or cis-acting regulatory variants near *G6PD* (promoter, enhancer, repressor, or insulator regions) were associated with G6PD activity, we repeated the association analysis described in the XWAS for all WGS variants present within one million base pairs of the *G6PD* CDS boundary. This *G6PD*-WAS analysis tested 11,682 variants, restricted to variants with over 90% genotyping rate and present in 3 or more alleles, and identified strong association signals with G6PD activity in the *G6PD* gene region (Figure 2C) with the strongest association being for the common G6PD deficiency-causing c.202G>A variant, as expected. The c.1311C>T variant, which has been described in people with low G6PD activity, showed no association with G6PD activity in any of the association tests. To determine if we could identify any new variants that associated with G6PD deficiency, and not just variants of known effect or variants linked to them, we removed all individuals homozygous, hemizygous, or compound heterozygous for WHO class I/II/III or A/B variants and repeated the analysis. None of the variants within one million base pairs from the *G6PD* gene region are common enough or effectual enough on G6PD activity to remain significant after removing individuals with variants already known to contribute to G6PD deficiency (Figure 2D); thus, we did not find any evidence to support association of any new coding, intron, or regulatory variants with G6PD activity.

Using power simulations with the permuted significance threshold of 7.7×10^-17^ corresponding to Figure 2D, we empirically demonstrate that this analysis was ∼80% powered to detect associations with a correlation of 0.3 (r^2^=0.09) in variants with an allele frequency of 0.006, ∼80% powered to detect associations with a correlation of 0.5 (r^2^=0.25) for variants present in at least 3 alleles (allele frequency of 0.0015, the rarest variants included in the analysis), and was approximately 100% powered to detect associations with a correlation of 0.4 (r^2^=0.16) in variants with an allele frequency of 0.0125 and higher. These simulated values are approximate since each allele frequency is represented by differing proportions of heterozygotes and homo/hemizygotes based on the actual data. For comparison to Figure 2C (which had a slightly larger sample size), the c.202G>A variant has a correlation of -0.39 and an allele frequency of 0.044, and the c.563C>T variant had a correlation of -0.3 and an allele frequency of 0.0015 and were both found to be statistically significant by the *G6PD*-WAS. **Supplemental Figure S4** shows the results of the simulation analysis across the continuum of allele frequencies and effect sizes for both the permuted p-value of 7.7×10^-17^ and the Bonferroni significance threshold, demonstrating that at a set sample size and phenotype distribution, allele frequency has less influence on power compared to the effect size being detected. These results quantify the capabilities of this association analysis by defining the allele frequency and effect size combinations that the analysis was powered to detect if the associations existed.

### Characterization of Nonsynonymous G6PD Genotype Combinations by Activity

The association studies are limited in their ability to detect variants that are too rare to be reliably tested by association analysis and limited by the complexity arising from haplotype structures that are important to G6PD activity and the presence of G6PD deficiency. To address this, we qualitatively analyzed the haplotype and diplotype structure of *G6PD* variants by creating combined genotype groups for all nonsynonymous SNPs found in the *G6PD* CDS for 1,255 individuals with interpretable G6PD activity data, WGS, and no missing nonsynonymous *G6PD* genotypes. Figure 3 shows the results of the G6PD activity for each genotype group (which can be interpreted as haplotypes) with individuals having a diagnosis of G6PD deficiency outlined by a solid black border around the data point. There were an additional 57 individuals with a diagnosis of G6PD deficiency who did not have activity data and thus are not shown in Figure 3; their genotype combinations for nonsynonymous variants in the *G6PD* CDS are included in **Table 1** along with the individuals with G6PD deficiency from Figure 3.

**Figure 3:**
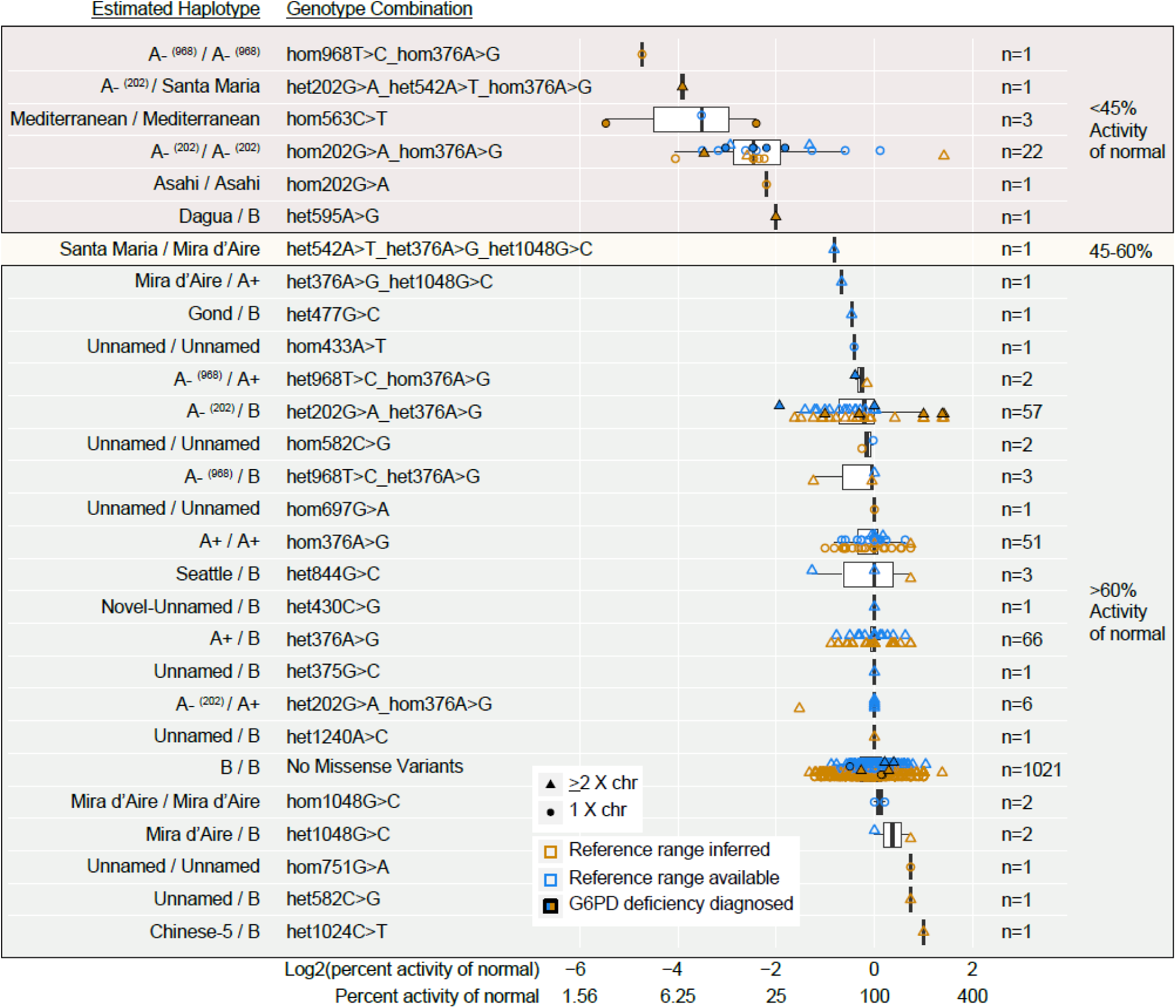
G6PD activity assay results for all unique missense genotype combinations seen in the All of Us v7.1 data for 1,255 individuals with G6PD assay interpretable results and 100% genotyping rate for *G6PD* CDS missense variants. The X-axis is scaled by Log2 transformed units to appropriately display the distance of values above and below 0 100%), with percent activity of normal also labeled. The Y-axis provides genotype combinations, estimated haplotype, and sample size for each group. The shaded boxes correspond to the median activity groupings using 45% and 60% as thresholds, based on the 2022 WHO guidance. Note: “hom” refers to both hemizygotes and homozygotes, and “het” refers to heterozygotes.

As expected, the WHO class IV or C missense variants (c.376A>G and c.1048G>C) did not confer loss of G6PD activity, though some individuals homozygous or heterozygous for c.376A>G were diagnosed with G6PD deficiency (**Supplemental Table S3 and Table 1**), which has been previously reported in rare cases.^40–42^ We identified six genotype combinations (haplotypes) for six variants with less than 45% normal activity (Figure 3), the cutoff used for WHO class B. One genotype combination had a median activity in the range of 45-60% which would be considered uncertain (WHO class U). Of the genotype combinations in individuals with median activity below 45%, five of these have previously been associated with decreased G6PD activity and hemolytic anemia, classified as Class II or III (c. 202G>A, c.[202G>A;376A>G], c.[376A>G;542A>T], c.[376A>G;968T>C], and c.563C>T). The sixth genotype that was <45% consists of the c.595A>G variant and has been previously reported in two individuals (at least one was hemizygous) with G6PD deficiency but without activity measurements, so it was a variant of uncertain significance (VUS).^43,44^ In All of Us the c.595A>G variant is seen in a heterozygous individual with 25% of the inferred normal activity and a diagnosis of G6PD deficiency. While most individuals that are heterozygous for G6PD deficiency-causing variants have normal activity (see other heterozygous haplotypes in Figure 3), some can be affected and thus there is now more evidence supporting that the c.595A>G variant can lead to G6PD deficiency.

Analysis of G6PD activity by genotype combinations (Figure 3) also supports that five missense VUS (c.433A>T, c.582C>G, c.697G>A, c.751G>A, and c.1048G>C), which are unclassified or have conflicting interpretations on ClinVar, may not be deleterious to G6PD since they were identified in separate hemizygous individuals without diagnosis of G6PD deficiency and with greater than 75% G6PD activity, above the threshold of 60% for nondeficiency.^16^ Each of these variants were also identified in additional individuals without data on G6PD activity or any record of G6PD deficiency diagnosed (**Supplemental Table S3**). Many other groups have classified c.1048G>C (G6PD Mira d’Aire) as unlikely to lead to G6PD deficiency and in Class IV,^45^ which our data support, though there are conflicting interpretations on ClinVar (Variation ID 93489). One variant, c.430C>G, has not been described before and was identified in one heterozygous individual. Although their G6PD activity is normal, this could be due to a sufficient fraction of their red blood cells expressing the B allele.

### Synonymous and Noncoding Variation Association with G6PD Activity

Most research on G6PD variants has focused on nonsynonymous variation, but with this rich dataset we also explored the effects of synonymous and noncoding variants. We analyzed all synonymous variant genotype combinations found in the *G6PD* CDS for 1,262 individuals with interpretable G6PD activity data, WGS, and no missing synonymous *G6PD* genotypes (**Supplemental Figure S5** shows these results in a format similar to Figure 3). The c.1116G>A, c.1311C>T, and c.1431C>T synonymous variants are observed in three or more hemizygous individuals and have a median activity over 60% of normal, the cutoff for WHO class C. This supports the current classification of c.1116G>A and enables us to classify c.1311C>T and c.1431C>T as WHO class C. This further supports their current classifications on ClinVar as benign/likely benign (variation IDs 10365, 470162, and 93496).

We further analyzed intron variants as potential causes for low G6PD activity in individuals without known genetic causes for low G6PD activity (those who are not homozygous or hemizygous for WHO class I/II/III or A/B variants) since the *G6PD*-WAS was not powered to evaluate rare genotypes. We identified 130 intron or UTR variants in at least one homozygous or hemizygous individual. Among these 130 variants, 58 were too rare to be tested in the *G6PD*-WAS. Of these 58 variants, statistical (**Supplemental Table S10**) and visual (**Supplemental Figure S6**) analysis revealed little evidence that any of these variants contribute to G6PD deficiency. Additionally, the total number of homozygous or hemizygous intron or UTR variants carried by an individual (ranging from 0 to 16) did not associate with the G6PD activity (p-value 0.16 by Pearson correlation test). Thus, it is unlikely that intron or UTR variants in *G6PD* contribute to low G6PD activity based on this dataset.

### Functional Characterization of G6PD Missense Variants of Uncertain Significance

To generate orthogonal evidence for classifying variants by their effect on G6PD, we chose to functionally test variants that were either newly identified, VUS in individuals with low G6PD activity, or VUS in hemi-or homozygous individuals with normal activity in the All of Us data. We measured the function of the newly identified c.430C>G variant and five other missense VUS (c.433A>T, c.582C>G, c.595A>G, c.697G>A, c.751G>A) by conducting functional assays in *Saccharomyces cerevisiae* yeast. For comparison, we also measured the function of variants established to have normal activity (c.376A>G, c.337G>A), decreased activity (c.[202G>A;376A>G], c.563C>T), and one variant associated with severe deficiency with chronic anemia (c.592C>T). Human *G6PD* can functionally complement the *S. cerevisiae* homolog *ZWF1* and restore the ability of yeast lacking *ZWF1* (*zwf1Δ*) to produce sufficient NADPH for synthesis of methionine and glutathione. Fully active G6PD variants enable growth of *zwf1Δ* yeast in the absence of methionine and presence of oxidants, but reduced function variants lead to slower growth.^46^ We expressed each variant in *zwf1Δ* yeast and measured growth in the absence of methionine and presence of hydrogen peroxide (**Supplemental Figure S7, Supplemental Table S5 and S11**). Yeast lacking *G6PD* grew significantly worse than ones expressing the normal-activity B allele, as did variants known to contribute to G6PD deficiency previously reported to have 1%-25% activity (A-(202), Mediterranean, Coimbra) (Figure 4).^10^ Also as expected, *G6PD* variants within the normal range of activity (A+, Sao Borja) grew similarly to yeast expressing the B allele.

**Figure 4:**
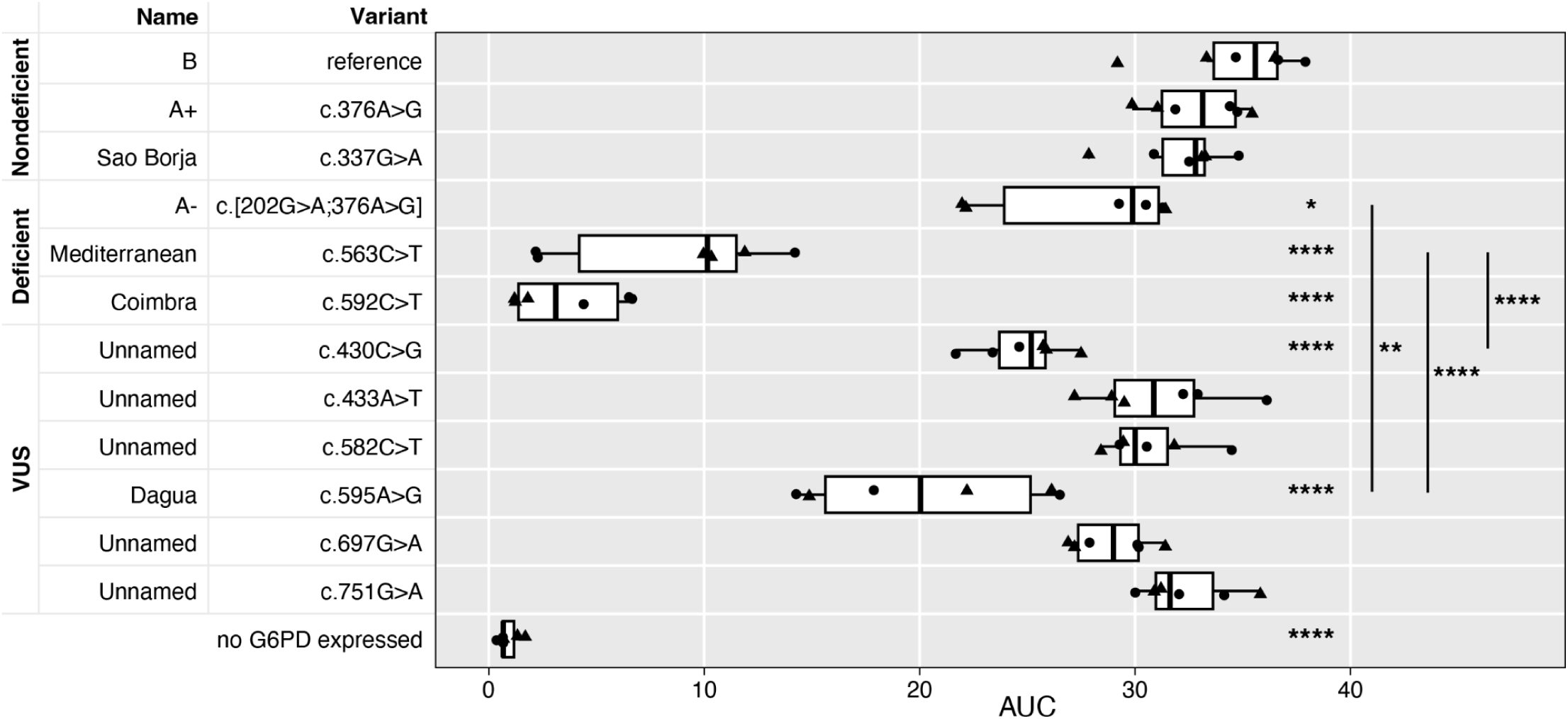
G6PD functional assay in *S. cerevisiae*. Growth (area under the curve) of *zwf1Δ S. cerevisiae* expressing indicated *G6PD* alleles in media lacking uracil and methionine with 1mM hydrogen peroxide. Shapes indicate biological replicates (independent transformants). Significance from reference unless otherwise indicated, by ANOVA with Tukey’s HSD; *, p<0.05; **, p<0.01; ****, p<0.0001.

Of the VUS, G6PD Dagua (c.595A>G) and c.430C>G led to reduced growth, while c.433A>T, c.582C>G, c.697G>A, and c.751G>A did not significantly reduce growth (p>0.05). This was consistent with activity measurements in red blood cells (Figure 3). However, since the growth of VUS without significantly reduced activity is also not significantly different from G6PD A-(202), with these conditions our assay cannot be used as evidence that variants do not contribute to deficiency.

Based on comparisons to variants of known effect, G6PD Dagua is significantly different from G6PD Mediterranean and A-(202), indicating an activity between 8-25%; yeast expressing c.430C>G grew significantly better than G6PD Mediterranean but not significantly worse than G6PD A-(202).

### Computational Prediction of Variant Effect

We also applied computational tools according to ClinGen recommendations^31^ for these VUS, and all variants listed in **Supplemental Table S3** to include variants of known effect as controls (**Supplemental Table S12**). Variants annotated in ClinVar as Benign or Likely Benign (c.311G>A, c.375G>C, c. 376A>G) and the WHO class C variant c.376A>G were predicted to be benign, and variants annotated in ClinVar as pathogenic (c.1360C>T, c.1388G>A) were predicted to be pathogenic, supporting our method for taking a consensus score from the tools applied (see Methods). However, the computational predictors did not unanimously identify c.563C>T, c.542A>T, and c.202G>A as pathogenic despite being annotated in ClinVar as pathogenic, indicating that some challenges remain in applying these tools to G6PD variants. The VUS of interest, c.433A>T, c.582C>G, and c.697G>A were predicted to be unlikely to disrupt G6PD function, and c.430C>G, c.595A>G, and c.751G>A were predicted to disrupt function (**Supplemental Table S12**). This generally agreed with our activity measurements from All of Us and our yeast assay, except for c.751G>A. This was similar to what we observed for c.1048G>C, which was computationally predicted to be pathogenic, but which we observed in two hemizygotes with normal activity (Figure 3) and has been previously reported to not lead to G6PD deficiency.^45^

### Variant Interpretation Updates

By applying ACMG guidelines for interpreting sequence variants^17^ to phenotype information from All of Us, our functional data, computational predictors,^31^ and previously published information, we interpret G6PD Dagua (c.595A>G) as pathogenic and c.430C>G as likely pathogenic. We interpret c.433A>T, c.582C>G, and c.697G>A as likely benign. We also provide additional pieces of evidence that could help with future interpretation of two other VUS (c.751G>A and c.1048G>C) which we observed in hemizygotes with normal activity but were computationally predicted to have reduced function and thus remain VUS due to conflicting and insufficient evidence. We also have evidence to support new or changed WHO classification of two synonymous variants (c.1311C>T and c.1431C>T) for a total of 9 new or changed G6PD variant interpretations (**Table 2**).

**Table 2:** Summary of added evidence and updated G6PD variant interpretations.

| CDS Variant | Previous Interpretation (Geck et al 2023) | ClinVar (March 2024) | Evidence | Suggested Interpretation Update |
| --- | --- | --- | --- | --- |
| c.430C>G | <u>WHO (2022):</u> NA<br><u>WHO (1985):</u> NA<br><u>ACMG:</u> NA | NA | <u>This Study:</u> Observed in a heterozygote with normal activity (**Figure 3**). Reduced activity in functional assay (**Figure 4**) (**PS3**), predicted to have decreased function (**PP3_M**) (**Supplemental Table S12**). Not found in gnomAD (**PM2**). | <u>WHO (2022):</u> **U**<br><u>ACMG:</u> **Likely pathogenic** |
| c.433A>T | <u>WHO (2022):</u> NA<br><u>WHO (1985):</u> Unknown<br><u>ACMG:</u> NA | Uncertain | <u>This Study:</u> Observed in a hemizygote with normal activity in red blood cells (**Figure 3**) (**BS3**). Predicted to have normal function (**Supplemental Table S12**) (**BP4**). | <u>WHO (2022):</u> **U**<br><u>ACMG:</u> **Likely benign** |
| c.582C>G | <u>WHO (2022):</u> NA<br><u>WHO (1985):</u> Unknown<br><u>ACMG:</u> Likely pathogenic | Conflicting | <u>This Study:</u> Observed in 2 hemizygotes with normal activity in red blood cells (**Figure 3**, one with normal range inferred) (**BS3**). Predicted to have normal function (**Supplemental Table S12**) (**BP4_M**).<br><u>Existing:</u> Observed in heterozygote with 9.2U/gHb activity in hemolysate, presumed deficient within range 4.6-9.8U/gHb, but is 70% of normal (13.2U/gHb),<sup>47</sup> which is widely considered non-deficient.<sup>16</sup> | <u>WHO (2022):</u> **U**<br><u>ACMG:</u> **Likely benign** |
| c.595A>G | <u>WHO (2022):</u> NA<br><u>WHO (1985):</u> Unknown<br><u>ACMG:</u> uncertain | Conflicting | <u>This Study:</u> Observed in a heterozygote with 25% normal activity (**Figure 3**) (**PP4**). Reduced activity in functional assay (**Figure 4**) (**PS3**). Predicted to have decreased function (**Supplemental Table S12**) (**PP3**). Located in substrate binding site (**PM1**) and very low frequency in gnomAD (**PM2**).<br><u>Existing:</u> Small reduction in functional assay (**PS3_M**),<sup>48</sup> and identified in two individuals (at least 1 hemizygote) with deficiency (**PP4**).<sup>43,44</sup> | <u>WHO (2022):</u> **U**<br><u>ACMG:</u> **Pathogenic** |
| c.697G>A | <u>WHO (2022):</u> NA<br><u>WHO (1985):</u> Unknown<br><u>ACMG:</u> NA | Uncertain | <u>This Study:</u> Observed in a hemizygote with normal activity in red blood cells (**Figure 3**, normal range inferred) (**BS3_P**). Predicted to have normal function (**Supplemental Table S12**) (**BP4**). | <u>WHO (2022):</u> **U**<br><u>ACMG:</u> **Likely benign** |
| c.751G>A | <u>WHO (2022):</u><br>NA<br><u>WHO (1985):</u><br>Unknown<br><u>ACMG:</u> NA | NA | <u>This Study:</u> Observed in a hemizygote with normal activity in red blood cells (**Figure 3**, normal range inferred) (**BS3_P**), but predicted to have reduced function (**Supplemental Table S12**) (**PP3**). | <u>WHO (2022):</u> **U**<br><u>ACMG:</u> **Uncertain** |
| c.1048G>C | <u>WHO (2022):</u><br>NA<br><u>WHO (1985):</u><br>IV<br><u>ACMG:</u> NA | NA | <u>This Study:</u> Observed in two hemizygotes with normal activity in red blood cells (**Figure 3**) (**BS3**), but predicted to have reduced function (**Supplemental Table S12**) (**PP3**).<br><u>Existing:</u> Observed in a hemizygote with deficiency but activity not stated (**PP4**).<sup>43</sup> | <u>WHO (2022):</u> **U**<br><u>ACMG:</u> **Uncertain** |
| c.1311C>T | <u>WHO (2022):</u><br>B<br><u>WHO (1985):</u><br>Synonymous<br><u>ACMG:</u><br>Uncertain | Benign | <u>This Study:</u> Observed in >3 hemi- and homozygotes with normal activity in red blood cells (**Supplemental Figure S5**) (**BS3**).<br><u>Existing:</u> Over 16% frequency in gnomAD (**BA1**). Identified in hemizygotes without deficiency (**BS2**).<sup>49,50</sup> Numerous other studies report activity; together with this study the weighted average activity is over 60% of normal (**BS3**). Additional interpretation on ClinVar support benign (**BP6**). | <u>WHO (2022):</u> **C**<br><u>ACMG:</u> **Benign** |
| c.1431C>T | <u>WHO (2022):</u><br>NA<br><u>WHO (1985):</u><br>Synonymous<br><u>ACMG:</u> NA | Benign/<br>Likely<br>benign | <u>This Study:</u> Observed in >3 hemi- and homozygotes with normal activity in red blood cells (**Supplemental Figure S5**) (**BS3**).<br><u>Existing:</u> Additional interpretation on ClinVar support benign (**BP6**). | <u>WHO (2022):</u> **C**<br><u>ACMG:</u> **Likely benign** |

## DISCUSSION

This study describes G6PD deficiency phenotypes in relation to variants found in *G6PD* in the All of Us Research Program v7.1 data. We identified 359 variants within the CDS of *G6PD* in 245,394 individuals. We provide evidence to interpret the variant of uncertain significance (VUS) *G6PD* c.595A>G (also called G6PD Dagua or Açores) as pathogenic for G6PD deficiency, and three missense VUS (c.433A>T, c.582C>G, c.697G>A) as likely benign, and have evidence that two other VUS (c.751G>A and c.1048G>C) do not decrease G6PD activity to below 45% of normal. Additionally, we report a variant, c.430C>G, in a heterozygous individual with normal G6PD activity, which has reduced activity in a functional assay and is likely pathogenic for G6PD deficiency. We also present evidence that synonymous variants c.1311C>T and c.1431C>T are benign/likely benign and WHO class C. The confirmation of known relationships between *G6PD* variants and G6PD deficiency supports the utility of All of Us biobank data for studying G6PD deficiency; furthermore, the ability to connect phenotypic data to rare *G6PD* variants emphasizes the value of performing sequencing on large patient cohorts with well curated clinical data. Our data also provide additional evidence to support testing for more variants within the CDS of *G6PD* to develop more inclusive pharmacogenetic genotyping panels.

This study relied on the clinical G6PD activity results in the All of Us Research Program records, demonstrating an effective process of inferring normal ranges for activity results. A challenge to the use of real-world evidence from a biobank is that it is inherently tied to uncontrolled variability involved in the documentation, collection, and interpretation of data that was originally intended for clinical benefit to one patient, not for scientific study. This includes the challenge of inferring the normal G6PD activity values when not provided by the testing lab, and that different G6PD activity assays are used by different institutions included in the All of Us Research Program data. We found that the LOINC standardization did not adequately enable like-for-like activity assay results to be grouped, requiring a custom manual process of inferring the normal ranges. Given that LOINC was intended to solve this problem by providing universal codes to identify medical tests and results,^51^ more effort in laboratory assay documentation could help improve analysis of this data. Despite this, our process of cleaning and curating real-world data to create a phenotype that can be used for association analyses can be used as a template for future similar analyses. Our report here demonstrates that this method results in expected genotype-phenotype assignments for variants of known function, supporting its utility for maximizing the amount of data that can be gained from biobank analysis. While we do not propose that inferring normal ranges is always feasible for clinical variant interpretation, it increases our ability to identify patterns in genotype-phenotype association, and we continue to encourage all testing labs to report their reference ranges along with results to enable better interpretation.

Boundaries of genetic regulatory regions are not always well established which makes identifying and predicting variants that could affect expression challenging. However, we did not identify variants outside the *G6PD* CDS +/-1 million base pairs from *G6PD* that were common enough or effectual enough to be significantly associated with G6PD activity. This finding was not surprising based on previous reports that variation outside of the CDS or in splice sites does not contribute to G6PD deficiency^5^ and suggests that the unexplained variability in G6PD activity for individuals without CDS variants likely comes from environmental or assay sources, which are established to contribute to variability of G6PD activity test results.^52–54^

We incidentally observed some interesting results from the permutation testing and power simulations that we did in support of the *G6PD* association analysis. The drastically higher permuted false positive threshold compared to Bonferroni should serve as a signal that permutation testing in GWAS analyses is a better method for distinguishing false positives, especially when sequencing data is used without LD pruning. Permutation testing has been described as the gold standard methodology for empirically determining the rate of false positives in GWAS,^25–27^ yet it is underutilized.

Many criticize the Bonferroni threshold as being too stringent because it penalizes each additional association test as if variants were completely independent; however, we found it to be too lenient. With the power simulations, we expected a more linear trend in statistical power increase as allele frequencies increased. However, we observed this effect only within a narrow range of effect sizes, and that variant effect sizes above or below this range were not greatly affected by allele frequency. Permutations and power simulations are similar to randomization procedures originally described by Fisher in 1935,^55^ and now are far more executable in the age of supercomputers, serving to provide empirical evidence towards the likelihood that observations originate from true differences or from random chance.

These tools helped us interpret the *G6PD* association analyses, leading to the conclusion that explaining additional interindividual variability in G6PD activity will need to come from analysis of rare variants, larger structural variants, or non-genetic sources.

This study finds that the prevalence of a documented diagnosis of G6PD deficiency in the All of Us Research Program cohort is vastly lower than the prevalence of genetic G6PD deficiency-causing genotypes, and below the global frequency of G6PD deficiency of approximately 5%.^4^ Among individuals homozygous or hemizygous for a WHO class I/II/III or A/B variant, over 99% do not have a documented G6PD deficiency diagnosis. This is not unexpected, firstly because many individuals would be asymptomatic unless they encountered a trigger. Since preemptive G6PD activity testing is only commonly done for a few drugs (e.g. rasburicase, pegloticase),^3^ asymptomatic individuals may never be diagnosed. While there is strong evidence that ingestion of fava beans or certain drugs are likely to reveal G6PD deficiency in treated individuals (dapsone, methylene blue, pegloticase, primaquine, rasburicase, tafenoquine, and toluene blue),^3^ only a subset of G6PD-deficient individuals would encounter these triggers. Additionally, in some cases self-reported ancestry may affect which individuals are recommended for G6PD testing, or which variants are genotyped, based on the frequency of *G6PD* variants in the region of their reported ancestry.^3^ The second reason that the frequency of G6PD deficiency is so low in All of Us may be that G6PD deficiency ICD codes are not always entered into health records, or G6PD deficiency could be managed outside systems that would document ICD codes. To this point, we observed many reported G6PD activity tests with low activity but no diagnosis of deficiency in the individual. This lack of reporting hampers the utility of the data for large-scale analysis of G6PD deficiency and associated phenotypes and is problematic for individuals who may be at risk for drug and food induced hemolytic anemia.

While G6PD deficiency often goes undiagnosed, the inverse is also true: at least 16 individuals with a diagnosis of G6PD deficiency in All of Us do not have a clear genetic basis for this disease (**Table 1**). Although there is a possibility of variant calling errors from the sequencing data, six of these diagnosed individuals had activity results as shown in Figure 3, and they were all normal. Since these individuals were reported with normal G6PD activity it is unclear if other hematological parameters or other unknown factors led to their diagnoses. This suggests the possibility that G6PD deficiency may be misdiagnosed, since by definition it leads to reduction of G6PD activity, and is well established to be a genetically driven trait. Our findings and others support that the incorporation of genetic testing into the differential diagnosis of suspected G6PD deficiency would result in more accurate diagnoses.^8^

A major challenge of implementing genetic testing for G6PD deficiency is the large number of variants. Many genetic testing panels for pharmacogene variants only include c.202G>A (or c.202G>A and c.376A>G), which, based on the All of Us population, would not identify 13% of affected individuals who carry other G6PD deficiency-causing variants.

Potential solutions to this problem include increasing the number of variants included on pharmacogenetic panels or by performing whole genome or exome sequencing in clinical scenarios where pharmacogenetic panels are insufficient.

Including *G6PD* genotype in the Medicine and Your DNA opt-in return-of-results to All of Us participants^9^ is an important step towards preemptive identification of individuals who would benefit from G6PD activity testing to determine if they have G6PD deficiency. To aid in this initiative and the general interpretation of these variants, we will submit the additional evidence for or against pathogenicity of the identified variants to ClinVar (**Table 2**). This evidence can also aid in classification of variants for returning genetic testing results to patients and caregivers. As more individuals with activity measurements for rare variants are identified to meet the classification requirement of measurement in three unrelated individuals, the WHO classifications can also be updated.^16^

The All of Us Research Program data coupled with our functional assay provided the necessary data to interpret two variants of previously uncertain significance. *G6PD* Dagua (c.595A>G) is very rare but has been described in three previous reports: two individuals (at least one was hemizygous) were reported with G6PD deficiency,^43,44^ and a small activity reduction was reported in a yeast-based growth-inhibition assay.^48^ The All of Us data provides additional clinical evidence of reduced activity, increasing the confidence that this variant is detrimental to G6PD activity. Additionally, we tested the variant using our own yeast-based growth-inhibition assay and found that it significantly reduces activity similar to variants well established to lead to G6PD deficiency. Using the same functional assay, we also provide evidence that c.430C>G, newly identified here in a heterozygote with normal G6PD activity, has reduced activity similar to the A-(202) variant. This indicates that, in a hemizygote or homozygote, it will likely lead to G6PD deficiency.

We identified five missense VUS in hemi- or homozygous individuals without a reduction in G6PD activity (c.433A>T, c. 582C>G, c.697G>A, c.751G>A, c.1048G>C), indicating that they are unlikely to lead to G6PD deficiency. All had been previously reported in gnomAD, and c.1048G>C was previously reported to ClinVar as likely benign by several groups. The c.1048G>C variant (G6PD Mira d’Aire) is classified as class IV by the 1985 WHO criteria but the evidence used to make this designation was not published.^45^ Therefore, our findings provide additional support that this variant does not lead to G6PD deficiency. However, due to conflicts with computational predictors and other existing evidence, we conservatively interpret this as uncertain until more evidence accumulates. Of the missense VUS of interest, only c.582C>G has previously reported activity data, in one heterozygote with slightly reduced activity categorized as “presumed deficient” but 70% of normal^47^ which is widely considered to be nondeficient.^16^ Our functional assay did not show significantly reduced activity for c.433A>T, c. 582C>G, c.697G>A, or c.751G>A, which is in agreement with the activity data from red blood cells. Interestingly, another variant at position 697 (c.697G>C), which leads to p.Val233Leu instead of p.Val233Ile, has been previously described as contributing to G6PD deficiency^56^ and has reduced activity when expressed in HEK293T cells.^57^ Overall, the results of our functional assay aligned with the reported activity or G6PD deficiency status from All of Us for homo- and hemizygous individuals, and were able to identify carriers of possible G6PD deficient variants.

In total, we have provided evidence to support new or updated classifications for nine variants according to both WHO and ACMG guidelines. The improvements to *G6PD* variant interpretation from this study would not be possible without the All of Us participants, whose contributions of genetic and electronic health record data enable a better understanding of which *G6PD* variants need to be considered for genetic tests. As future data is released by the All of Us Research Program and other diverse biobanks, it is likely that more *G6PD* variants will become interpretable, increasing the utility of genetic testing to identify G6PD-deficient individuals. We envision that our results will enable the design, and prompt increased use, of *G6PD* genetic testing strategies that test for all clinically relevant *G6PD* variants, including rare variants. Validating genetic tests as an appropriate means to diagnose G6PD deficiency will improve the accurate diagnosis of G6PD-deficient individuals and alleviate concerns about inaccurate point-in-time activity results. As the All of Us Research Program increases to one million participants, we expect that the combination of biobank analysis and functional assays will provide a more complete picture of which *G6PD* variants contribute to G6PD deficiency, enabling improved genotype interpretation to preemptively identify individuals at risk for adverse drug reactions.

## Search terms

G6PD, Glucose-6-phosphate dehydrogenase, All of Us, G6PD deficiency, LOINC, phenotype curation, pharmacogenetics, GWAS, permutation testing, power simulation, ACMG, WHO, ClinVar, ClinGen, variant interpretation, hemolytic anemia.

## Author contributions

Conceptualization of research (NRP, RCG, MD); Execution of All of Us primary analysis (NRP); Execution of functional yeast assays (RCG); curation of G6PD interpretation updates (RCG, NRP); Variant interpretation according to ACMG and WHO (RCG, NRP); Execution of computational predictors (RCG, NRP); Second person verification of All of Us results (TS, DL); Manuscript writing (NRP, RCG, TS, TCS, MD, DL). Funding of research (TCS, MD, NRP, RCG).

## Data and Resource Availability

The All of Us data was openly accessible at the initiation of this study (https://allofus.nih.gov/). All code used to generate the All of Us analyses is documented in the All of Us cloud computing platform as step-by-step notebooks in a dedicated workspace. We will share the workspace as read-only with approved users of the All of Us controlled tier data upon request. Code for analysis of *S. cerevisiae* functional assays are available at https://github.com/reneegeck/DunhamLab and strains are available upon request. All data produced in the present work are contained in the manuscript.

## Data Availability

All code used to generate the All of Us analyses is documented in the All of Us cloud computing platform as step-by-step notebooks in a dedicated workspace. We will share the workspace as read-only with approved users of the All of Us controlled tier data upon request. Code for analysis of S. cerevisiae functional assays are available at https://github.com/reneegeck/DunhamLab and strains are available upon request. All data produced in the present work are contained in the manuscript.

https://github.com/reneegeck/G6PDcat

https://github.com/reneegeck/DunhamLab

## Acknowledgements

We gratefully acknowledge *All of Us* participants for their contributions, without whom this research would not have been possible. We also thank the National Institutes of Health’s *All of Us* Research Program for making available the participant data examined in this study. An exception was granted by the All of Us Research Access Board to share data from groups of individuals below sample sizes of 20.

## SUPPLEMENTAL FIGURES AND TABLES

**Supplemental Figure S1:**
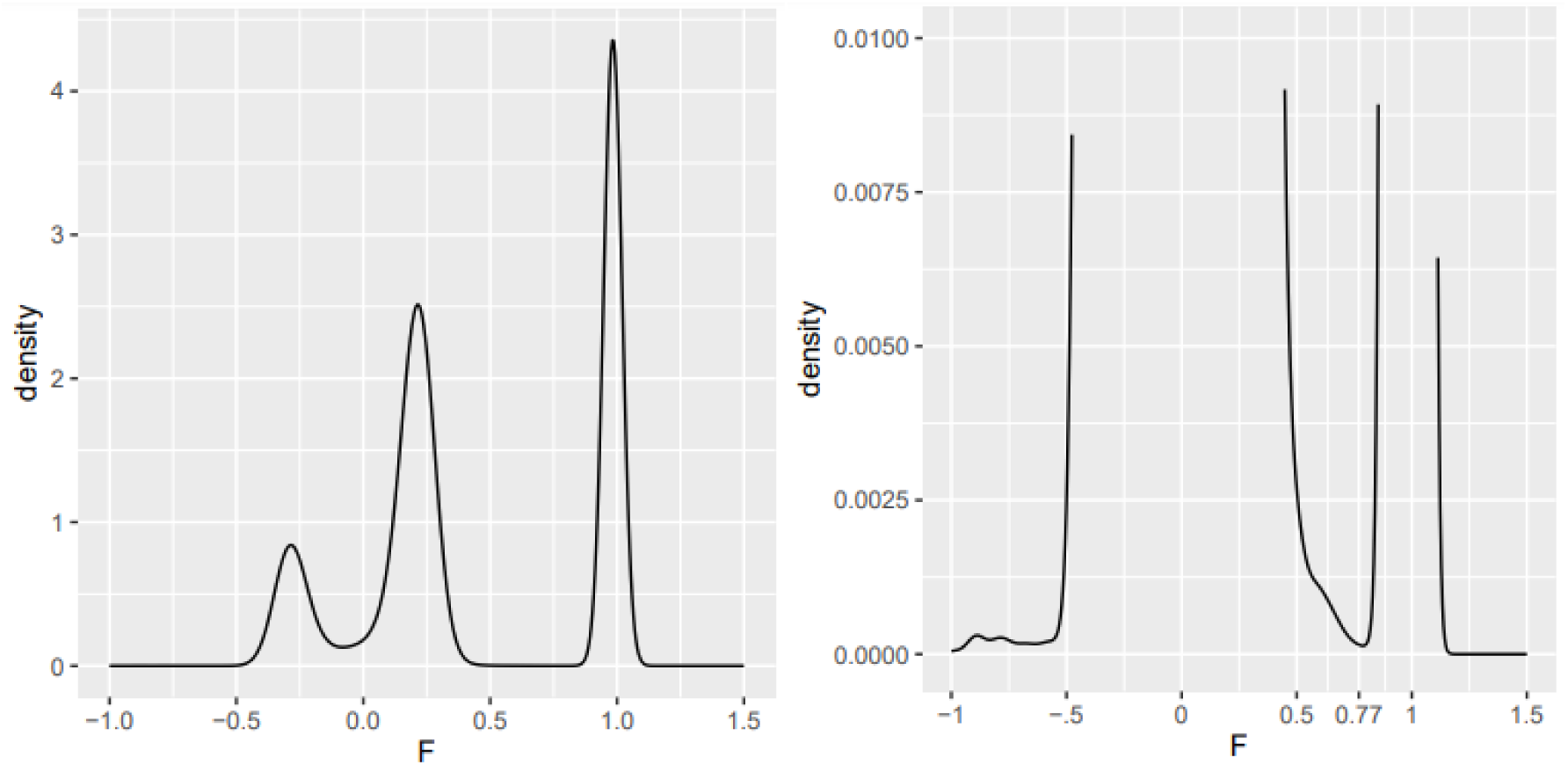
Imputed sex F coefficient (homozygosity rates) in the All of Us v7 participants with short read WGS. Left: full range. Right: zoomed in.

**Supplemental Figure S2:**
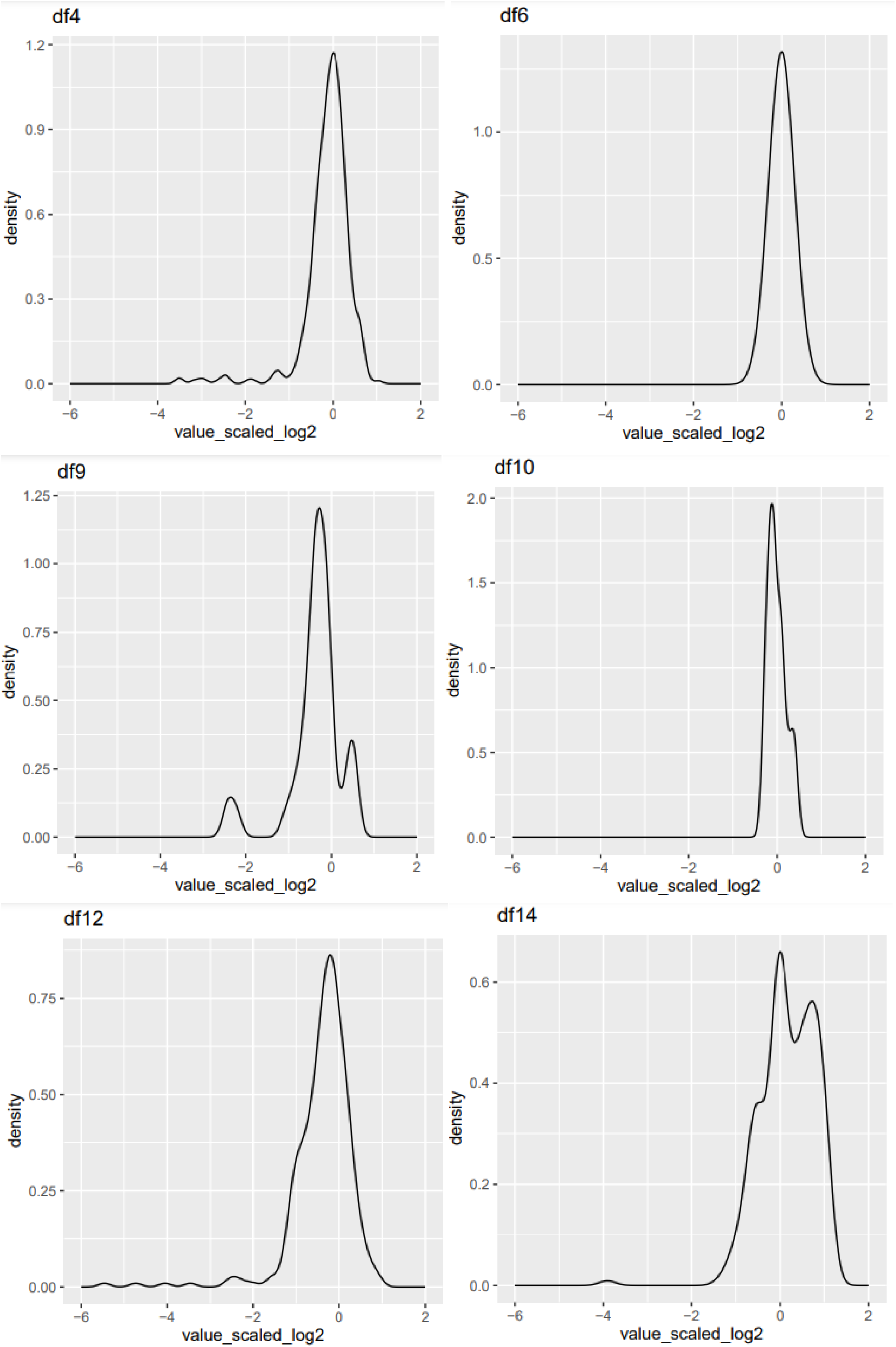
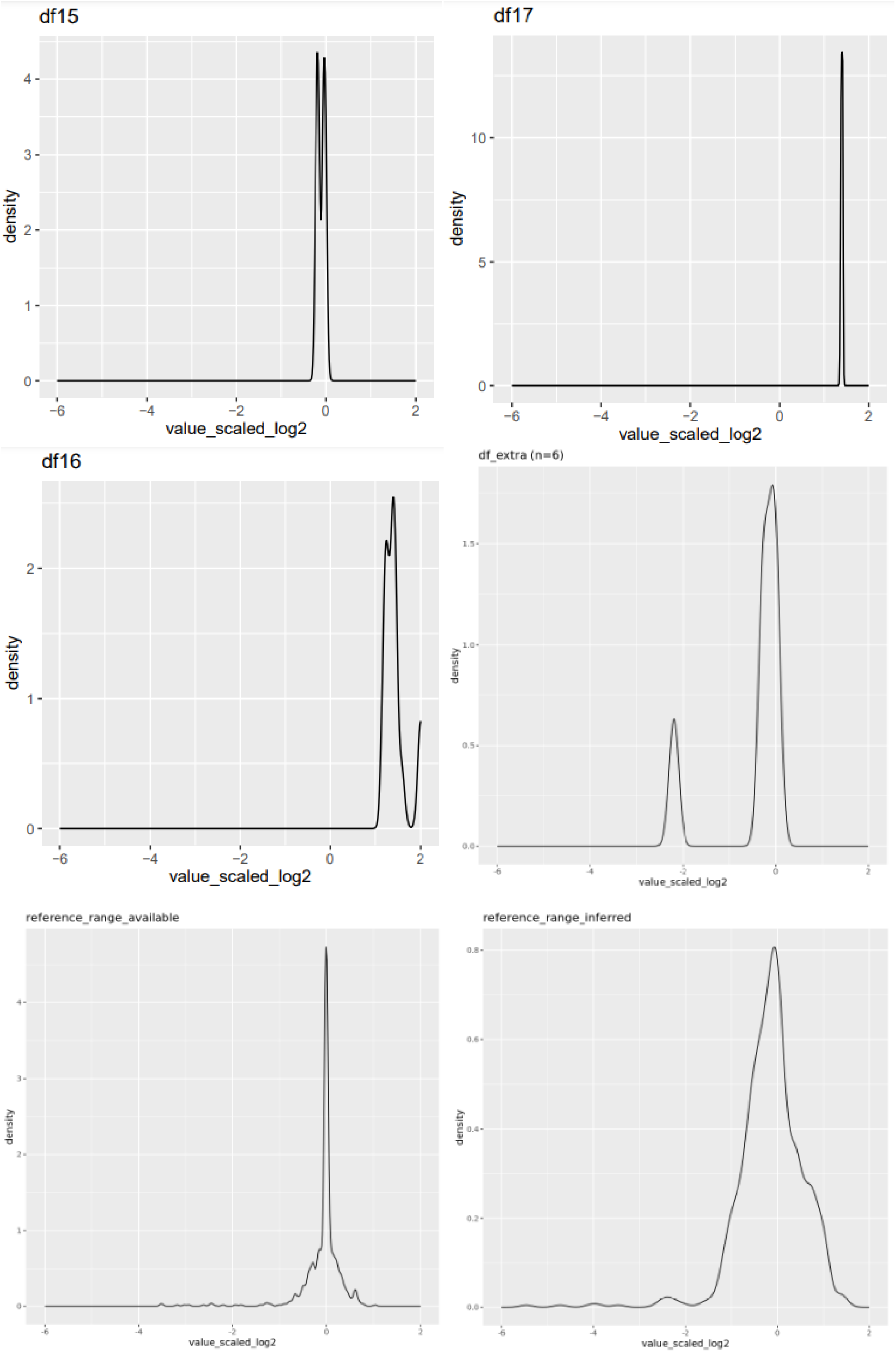
Distributions of each activity phenotype group corresponding to the data frames listed in **Figure 1** of the main manuscript. Default smoothing settings by ggplot2 geom_density were used. Additionally, the distributions are shown for the data when grouped by those with available reference ranges and those which were inferred.

**Supplemental Figure S3:**
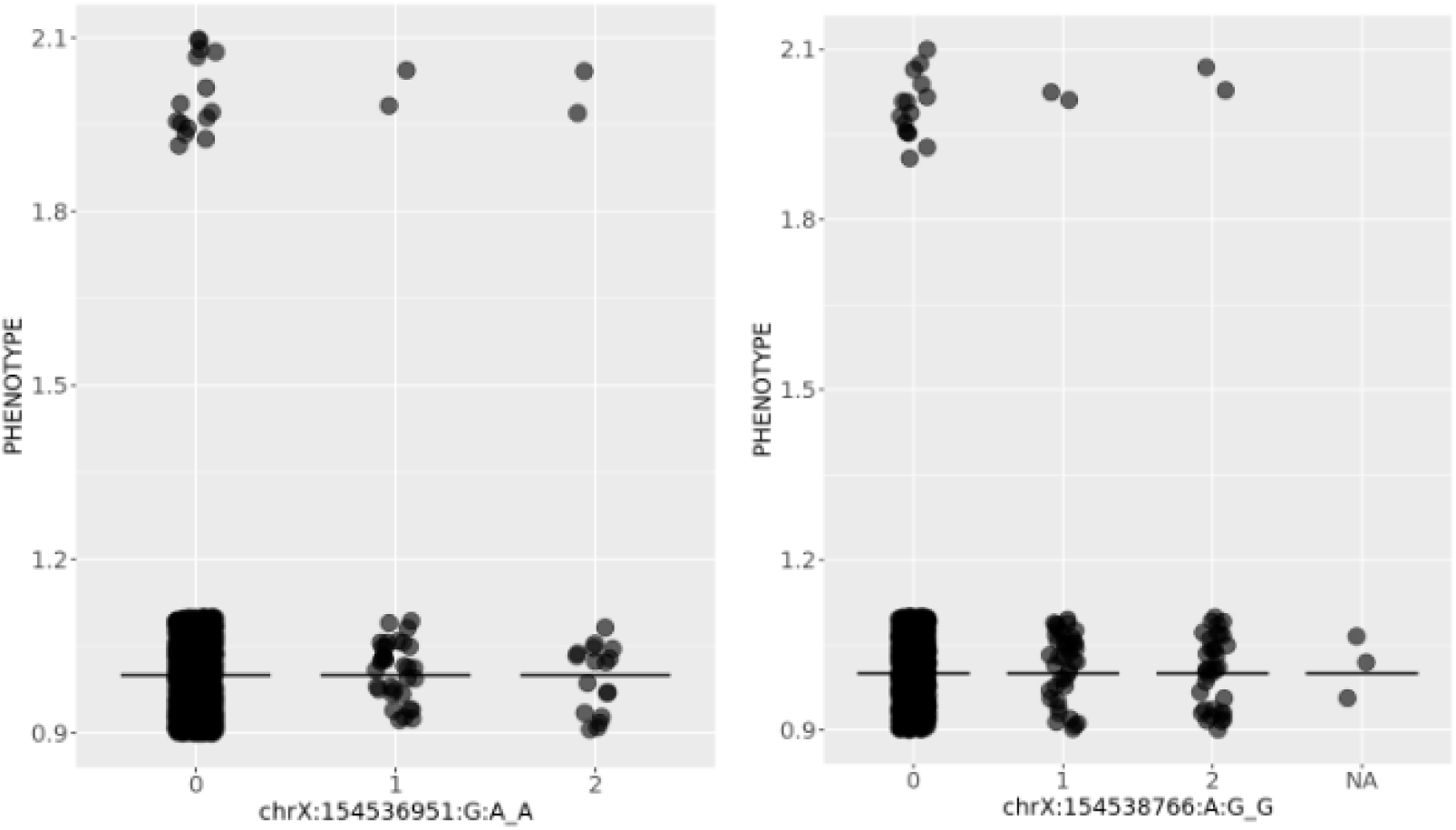
Visual assessment of the top 2 lowest p-value results from the Fisher exact enrichment analysis. Phenotype is coded as 1= no G6PD deficiency diagnosis, 2= G6PD deficiency diagnosis. Corresponds to Supplemental Table S9.

**Supplemental Figure S4:**
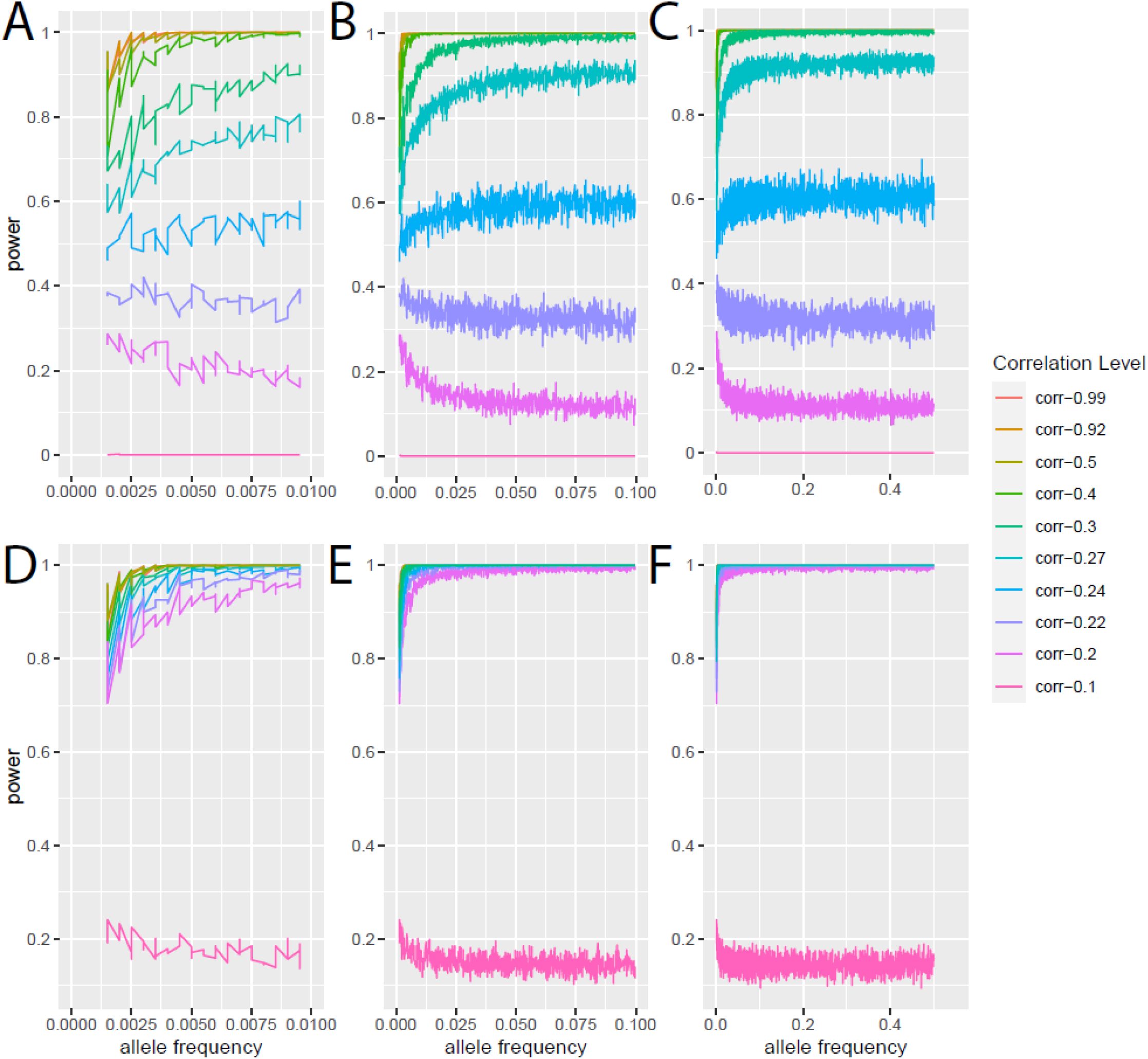
Power simulation corresponding to Figure 2D. A-C show the power simulations using an alpha of 7.7e-17 based on the permuted threshold, and D-F for the Bonferroni alpha. We found that increasing the sample size caused all correlations to have larger power as expected (not shown).

**Supplemental Figure S5:**
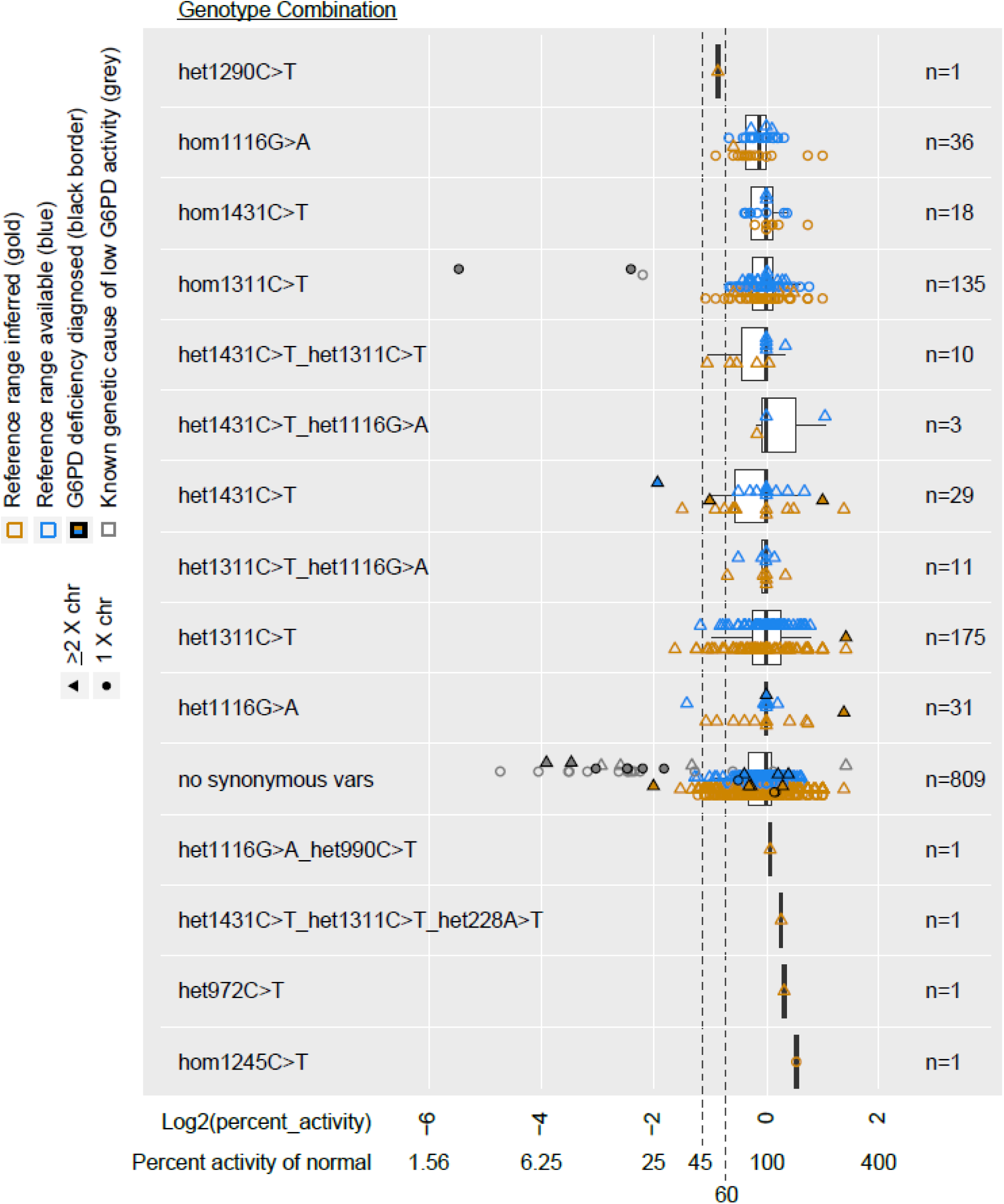
G6PD activity assay results for all unique synonymous genotype combinations seen in the All of Us v7.1 data for 1,262 individuals with G6PD assay interpretable results and 100% genotyping rate for G6PD CDS synonymous variants. The X-axis is Log2 transformed to appropriately display the distance of values above and below 0 (100%). The Y-axis provides genotype combinations, estimated haplotype, and sample size for each group. Note: “hom” refers to both hemizygotes and homozygotes.

**Supplemental Figure S6:**
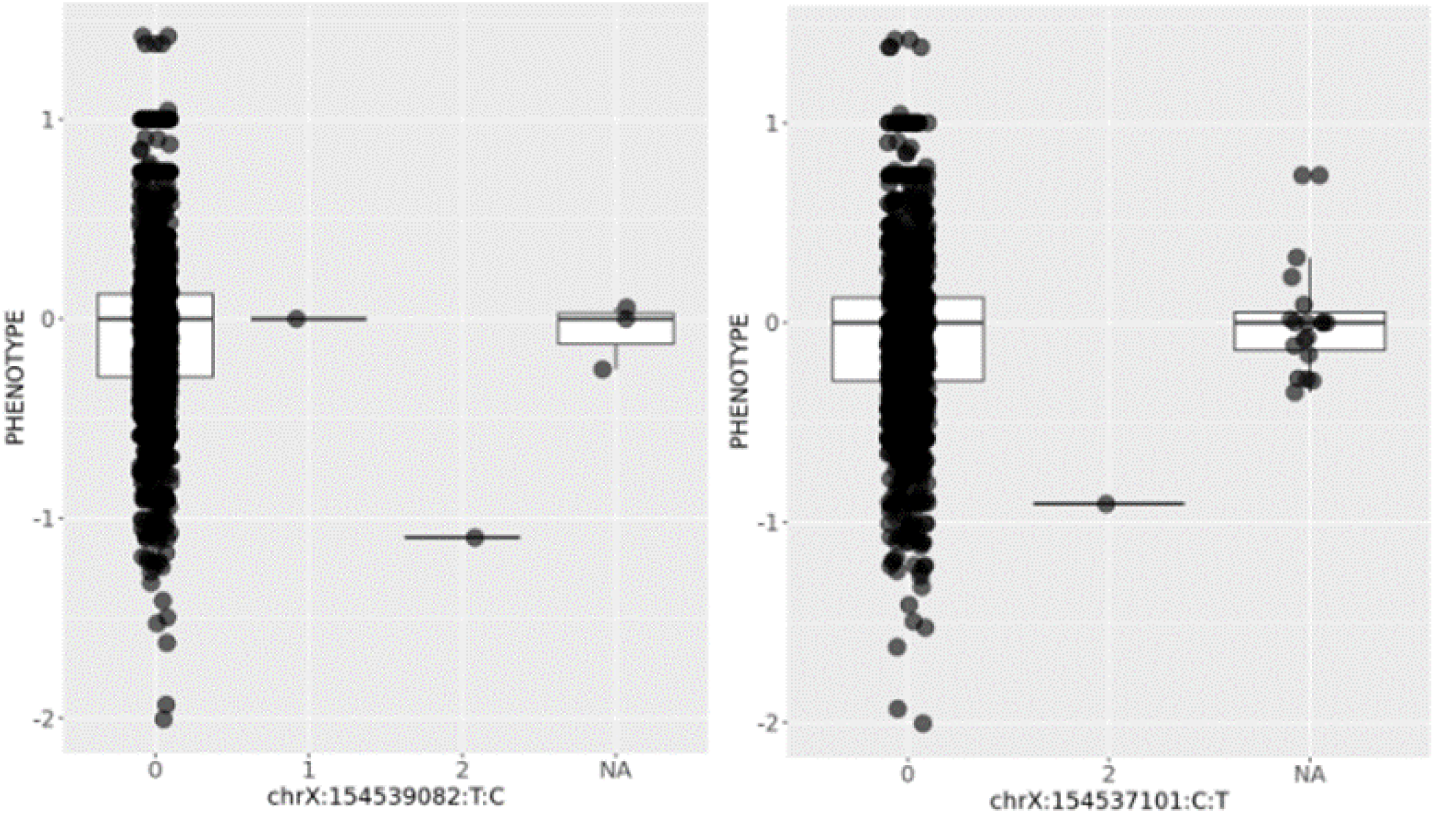
Visual assessment of the top 2 lowest p-value results from the Intron and UTR analysis among rare variants (maximum allele count = 2) and G6PD activity. Corresponds to Supplemental Table S10.

**Supplemental Figure S7:**
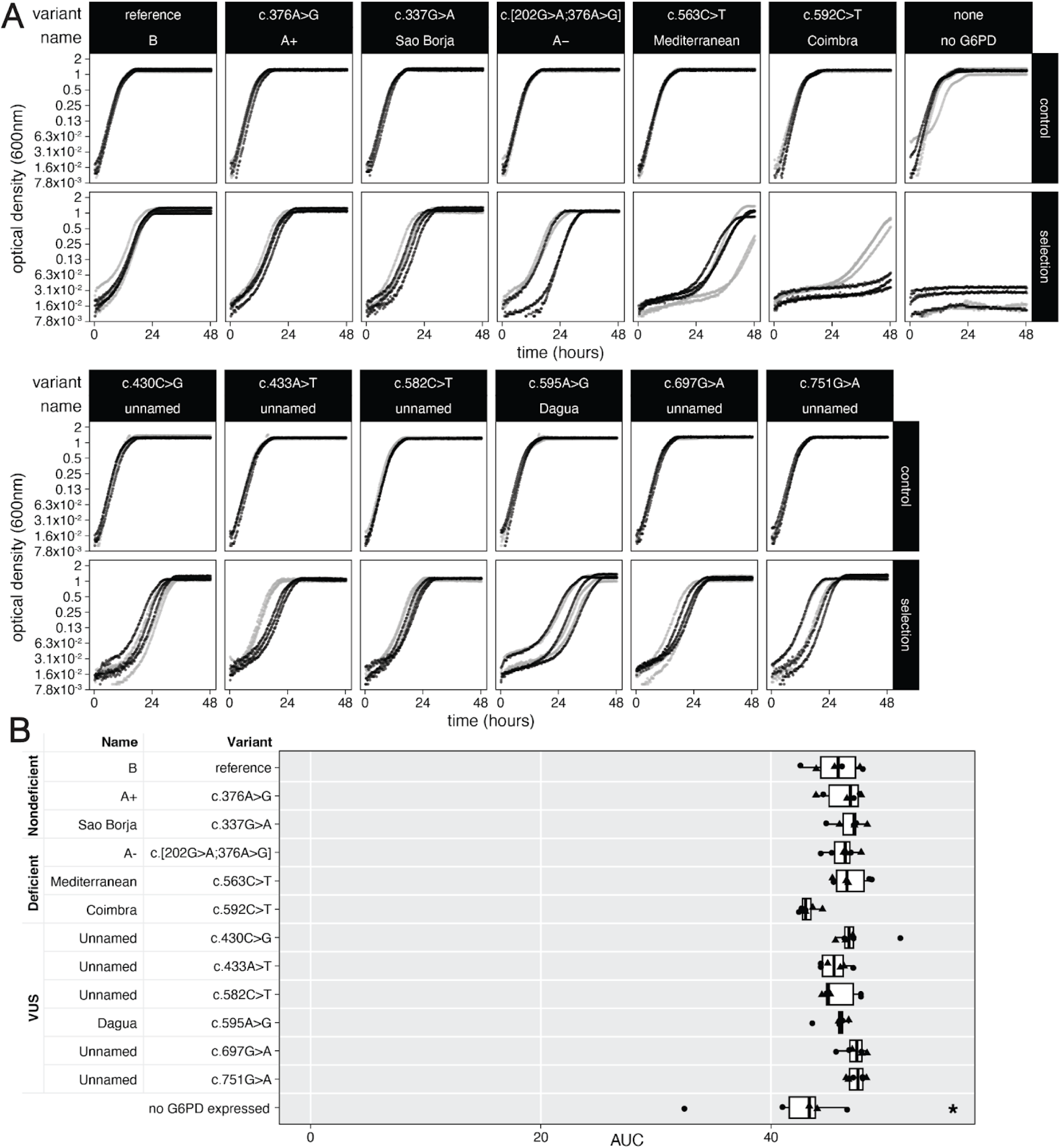
G6PD functional assay in *S. cerevisiae*. (A) Growth of *zwf1Δ S. cerevisiae* expressing indicated *G6PD* alleles in control media (c-ura) or selection for G6PD activity (c-ura-met +1mM hydrogen peroxide). Black and gray indicate the two different biological replicates (transformants). (B) Area under the curve for control media condition (c-ura) shown in A. Shapes indicate biological replicates (transformants); circles correspond with gray points in A. Significance from reference by ANOVA with Tukey’s HSD; *, p<0.05.

**Supplemental Table S1:** G6PD chromosomal coordinates that correspond to the canonical Ensembl protein coding sequence (CDS) coordinates used to filter Plink files. ENST00000393562 was used to define these regions.

| Chr | Start | End | Length |
| --- | --- | --- | --- |
| X | 154532000 | 154532090 | 91 |
| X | 154532188 | 154532280 | 93 |
| X | 154532386 | 154532462 | 77 |
| X | 154532567 | 154532802 | 236 |
| X | 154532942 | 154533128 | 187 |
| X | 154533576 | 154533669 | 94 |
| X | 154534035 | 154534160 | 126 |
| X | 154534338 | 154534496 | 159 |
| X | 154535168 | 154535385 | 218 |
| X | 154535937 | 154536045 | 109 |
| X | 154536141 | 154536178 | 38 |
| X | 154546036 | 154546155 | 120 |

**Supplemental Table S2:** Standard Concept Codes/Names for all laboratory tests with G6PD or Glucose-6-Phosphate dehydrogenase listed in the name.

| Standard Concept Code | Standard Concept Name (standard vocabulary = LOINC) | Count (rows of data) |
| --- | --- | --- |
| 32546-4 | Glucose-6-Phosphate dehydrogenase [Enzymatic activity/mass] in Red Blood Cells | 756 |
| 2357-2 | Glucose-6-Phosphate dehydrogenase [Enzymatic activity/volume] in Red Blood Cells | 410 |
| 2358-0 | Glucose-6-Phosphate dehydrogenase [Presence] in Serum | 296 |
| 21680-4* | G6PD gene mutations found [Identifier] in Blood or Tissue by Molecular genetics method Nominal | 240 |
| 2356-4 | Glucose-6-Phosphate dehydrogenase [Presence] in Red Blood Cells | 172 |
| 49502-8 | Glucose-6-Phosphate dehydrogenase [Entitic Catalytic Activity] in Blood | 64 |
| 2359-8 | Glucose-6-Phosphate dehydrogenase [Enzymatic activity/volume] in Serum | 18 |
| 33287-4 | Glucose-6-Phosphate dehydrogenase [Presence] in DBS | 16 |
|  | **Total** | **1,972** |
| 104692005^ | Glucose-6-phosphate dehydrogenase measurement^ | 38^ |

\*G6PD gene mutations test did not provide the results of the genetic test, only that it was performed
^This data was found during second person verification of the phenotypes and added to the final data.

**Supplemental Table S3.**
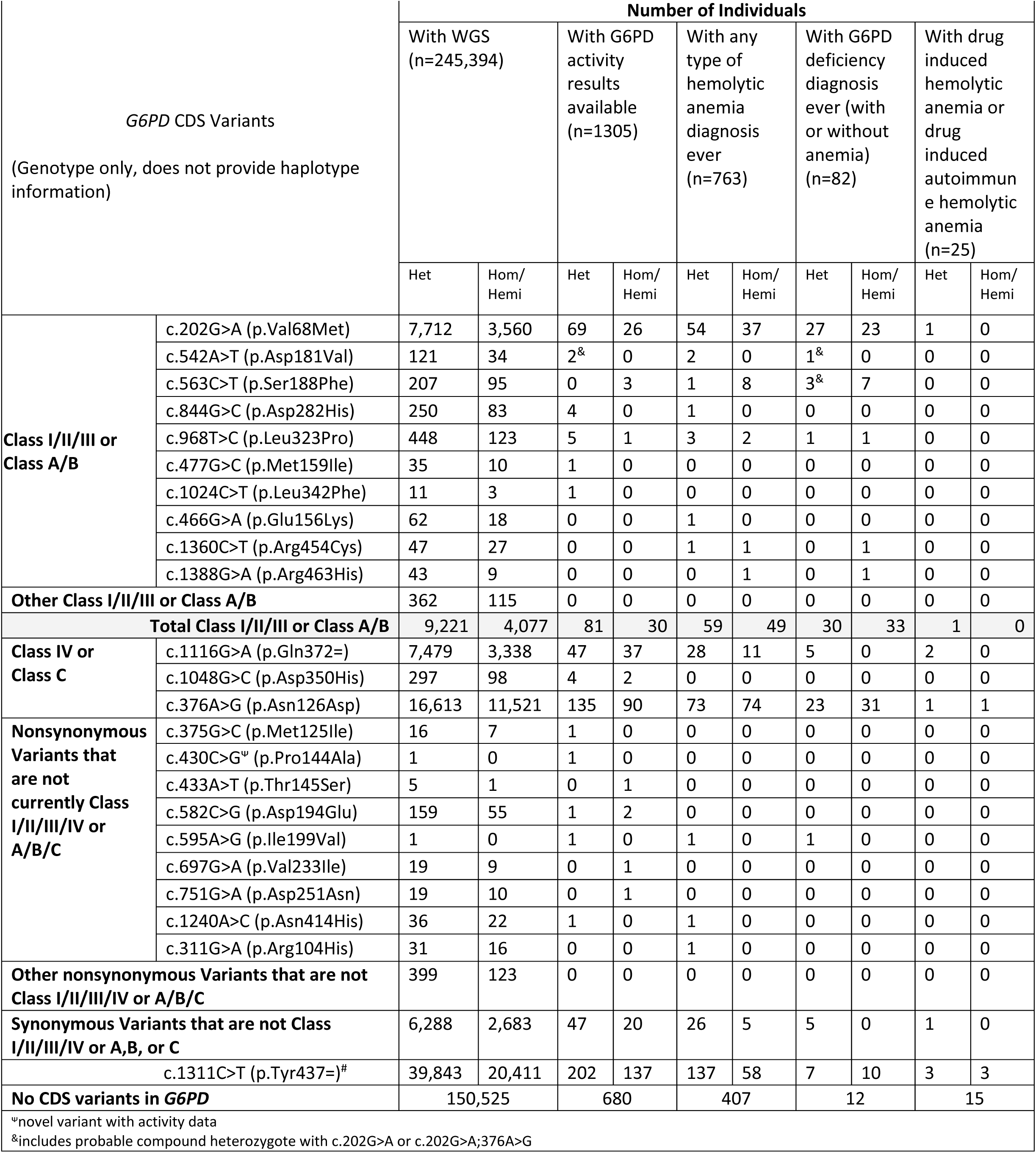
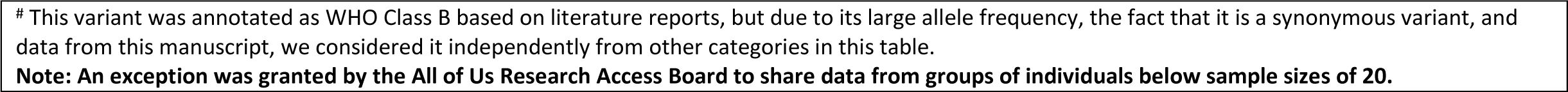
Unphased G6PD CDS variants found in the AoU individuals. Haplotypes are not provided here due to the uncertainty involved in phasing short read WGS.

**Supplemental Table S4:** Standard Concept Names for diagnoses for hemolytic anemia category

| Diagnosis standard concept name | Count (rows of data) |
| --- | --- |
| Autoimmune hemolytic anemia | 3279 |
| Acquired hemolytic anemia | 1223 |
| Hereditary spherocytosis | 763 |
| Hereditary hemolytic anemia | 706 |
| Hemolytic uremic syndrome | 668 |
| Non-autoimmune hemolytic anemia | 456 |
| Hemolytic anemia due to glutathione metabolism disorder | 408 |
| Glucose-6-phosphate dehydrogenase deficiency anemia | 280 |
| Evans syndrome | 224 |
| Hereditary elliptocytosis | 160 |
| Hemoglobinuria due to hemolysis from external causes | 90 |
| Cold autoimmune hemolytic anemia | 42 |
| Anemia due to enzyme deficiency | 40 |
| Hemolytic anemia due to drugs | 40 |
| Warm autoimmune hemolytic anemia | 37 |
| Atypical hemolytic uremic syndrome | 36 |
| Thrombotic thrombocytopenic purpura | 26 |
| Drug-induced autoimmune hemolytic anemia | 18 |
| Hemolytic disease of fetus OR newborn due to isoimmunization | 8 |
| Hemolytic disease of fetus OR newborn due to RhD isoimmunization | 7 |
| Hemolytic anemia | 5 |
| Hemolytic disease of fetus OR newborn due to ABO immunization | 2 |
| Acquired thrombotic thrombocytopenic purpura | 1 |
| Autoimmune hemolytic anemia mixed type | 1 |
| Secondary autoimmune hemolytic anemia co-occurrent and due to systemic lupus erythematosus | 1 |

**Supplemental Table S5:** *S. cerevisiae* strains used or created in this study

| Strain | G6PD allele | Genotype |
| --- | --- | --- |
| YMD4438 | none | *zwf1::KanMX ura3- HIS3 LEU2 LYS2* |
| YMD4439 | none | *zwf1::KanMX ura3- HIS3 LEU2 LYS2 [p416CYC1]* |
| YMD4441 | none | *zwf1::KanMX ura3- HIS3 LEU2 LYS2 [p416CYC1]* |
| YMD4440 | reference | *zwf1::KanMX ura3- HIS3 LEU2 LYS2 [p416CYC1-G6PD]* |
| YMD4442 | reference | *zwf1::KanMX ura3- HIS3 LEU2 LYS2 [p416CYC1-G6PD]* |
| YMD5009 | c.376A>G | *zwf1::KanMX ura3- HIS3 LEU2 LYS2 [p416CYC1-G6PD-A+]* |
| YMD5010 | c.376A>G | *zwf1::KanMX ura3- HIS3 LEU2 LYS2 [p416CYC1-G6PD-A+]* |
| YMD5011 | c.337G>A | *zwf1::KanMX ura3- HIS3 LEU2 LYS2 [p416CYC1-G6PD-SaoBorja]* |
| YMD5012 | c.337G>A | *zwf1::KanMX ura3- HIS3 LEU2 LYS2 [p416CYC1-G6PD-SaoBorja]* |
| YMD5013 | c.[202G>A;376A>G] | *zwf1::KanMX ura3- HIS3 LEU2 LYS2 [p416CYC1-G6PD-A-]* |
| YMD5014 | c.[202G>A;376A>G] | *zwf1::KanMX ura3- HIS3 LEU2 LYS2 [p416CYC1-G6PD-A-]* |
| YMD5015 | c.563C>T | *zwf1::KanMX ura3- HIS3 LEU2 LYS2 [p416CYC1-G6PD-Mediterranean]* |
| YMD5016 | c.563C>T | *zwf1::KanMX ura3- HIS3 LEU2 LYS2 [p416CYC1-G6PD-Mediterranean]* |
| YMD5017 | c.592C>T | *zwf1::KanMX ura3- HIS3 LEU2 LYS2 [p416CYC1-G6PD-Coimbra]* |
| YMD5018 | c.592C>T | *zwf1::KanMX ura3- HIS3 LEU2 LYS2 [p416CYC1-G6PD-Coimbra]* |
| YMD5019 | c.430C>G | *zwf1::KanMX ura3- HIS3 LEU2 LYS2 [p416CYC1-G6PD-430C>G]* |
| YMD5020 | c.430C>G | *zwf1::KanMX ura3- HIS3 LEU2 LYS2 [p416CYC1-G6PD-430C>G]* |
| YMD5021 | c.433A>T | *zwf1::KanMX ura3- HIS3 LEU2 LYS2 [p416CYC1-G6PD-433A>T]* |
| YMD5022 | c.433A>T | *zwf1::KanMX ura3- HIS3 LEU2 LYS2 [p416CYC1-G6PD-433A>T]* |
| YMD5023 | c.582C>T | *zwf1::KanMX ura3- HIS3 LEU2 LYS2 [p416CYC1-G6PD-582C>T]* |
| YMD5024 | c.582C>T | *zwf1::KanMX ura3- HIS3 LEU2 LYS2 [p416CYC1-G6PD-582C>T]* |
| YMD5025 | c.595A>G | *zwf1::KanMX ura3- HIS3 LEU2 LYS2 [p416CYC1-G6PD-Dagua]* |
| YMD5026 | c.595A>G | *zwf1::KanMX ura3- HIS3 LEU2 LYS2 [p416CYC1-G6PD-Dagua]* |
| YMD5027 | c.697G>A | *zwf1::KanMX ura3- HIS3 LEU2 LYS2 [p416CYC1-G6PD-697G>A]* |
| YMD5028 | c.697G>A | *zwf1::KanMX ura3- HIS3 LEU2 LYS2 [p416CYC1-G6PD-697G>A]* |
| YMD5029 | c.751G>A | *zwf1::KanMX ura3- HIS3 LEU2 LYS2 [p416CYC1-G6PD-751G>A]* |
| YMD5030 | c.751G>A | *zwf1::KanMX ura3- HIS3 LEU2 LYS2 [p416CYC1-G6PD-751G>A]* |

**Supplemental Table S6:**
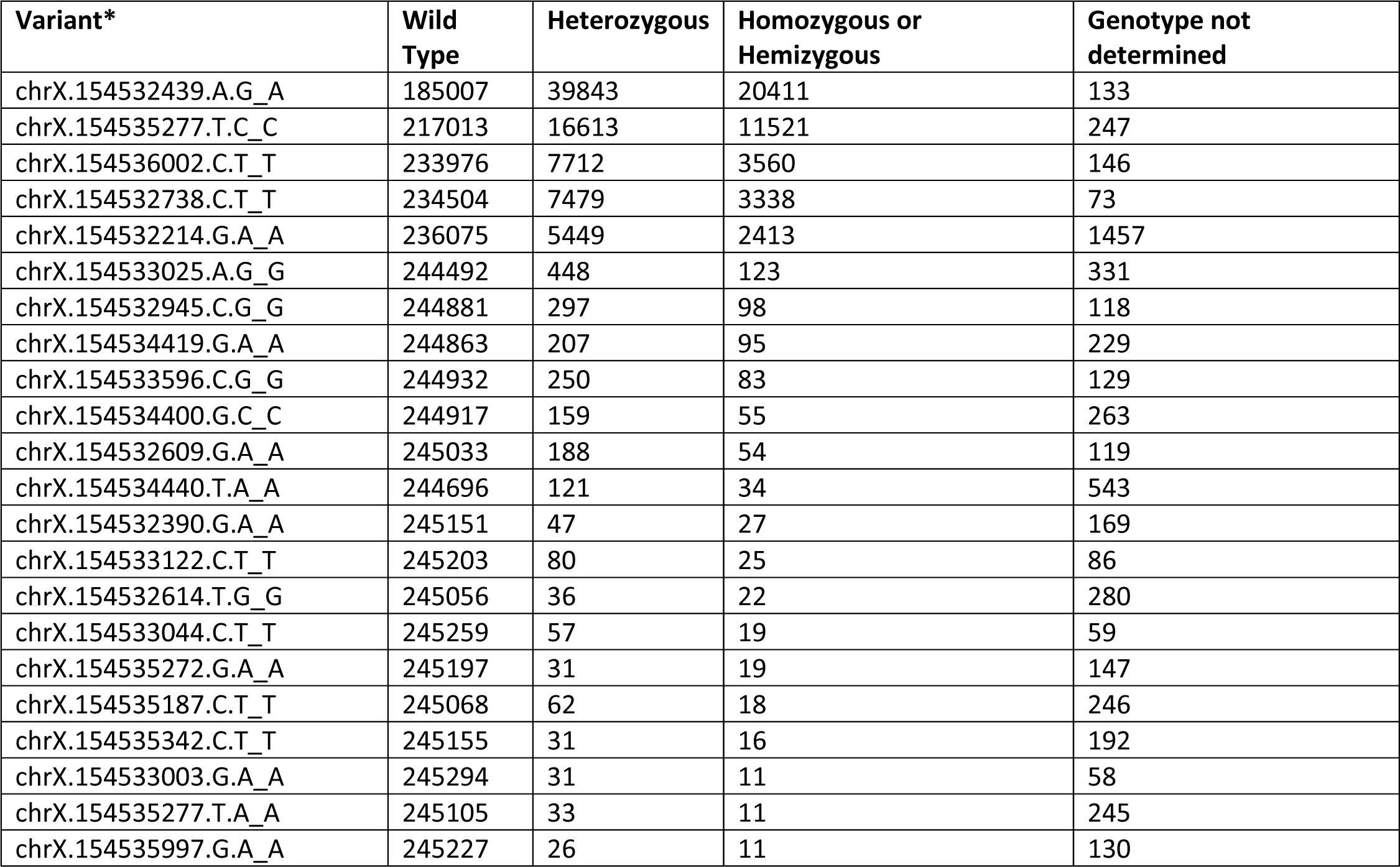
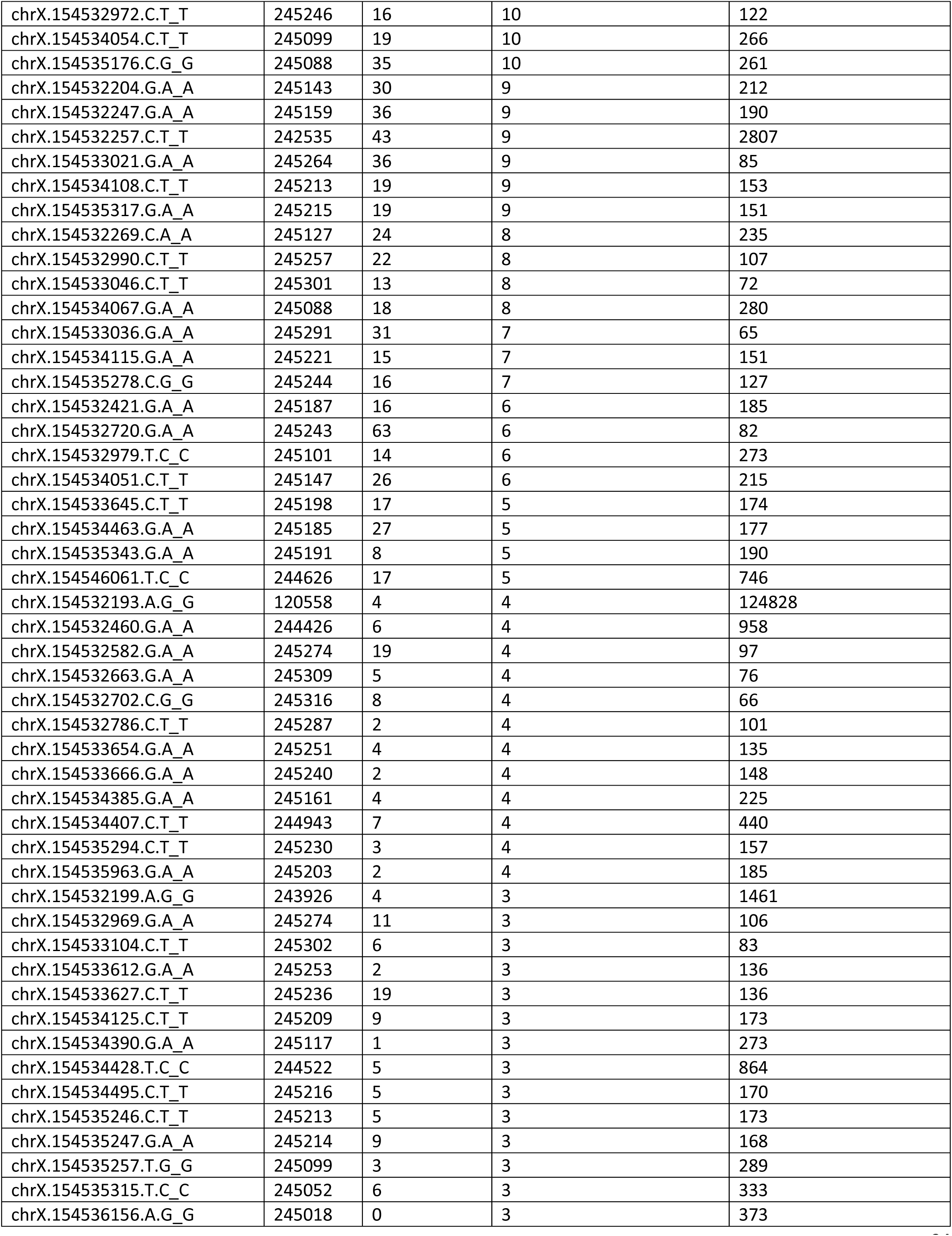
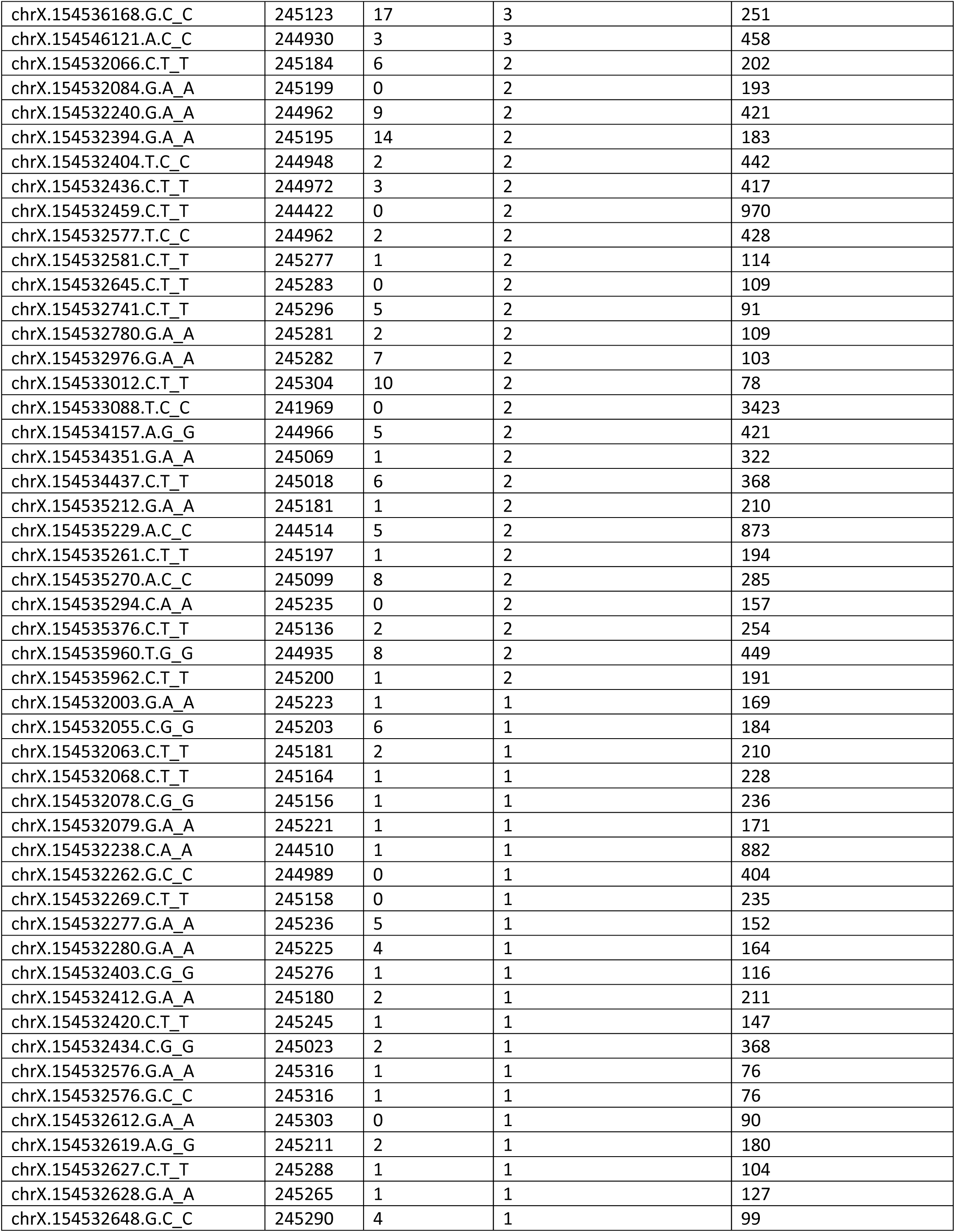
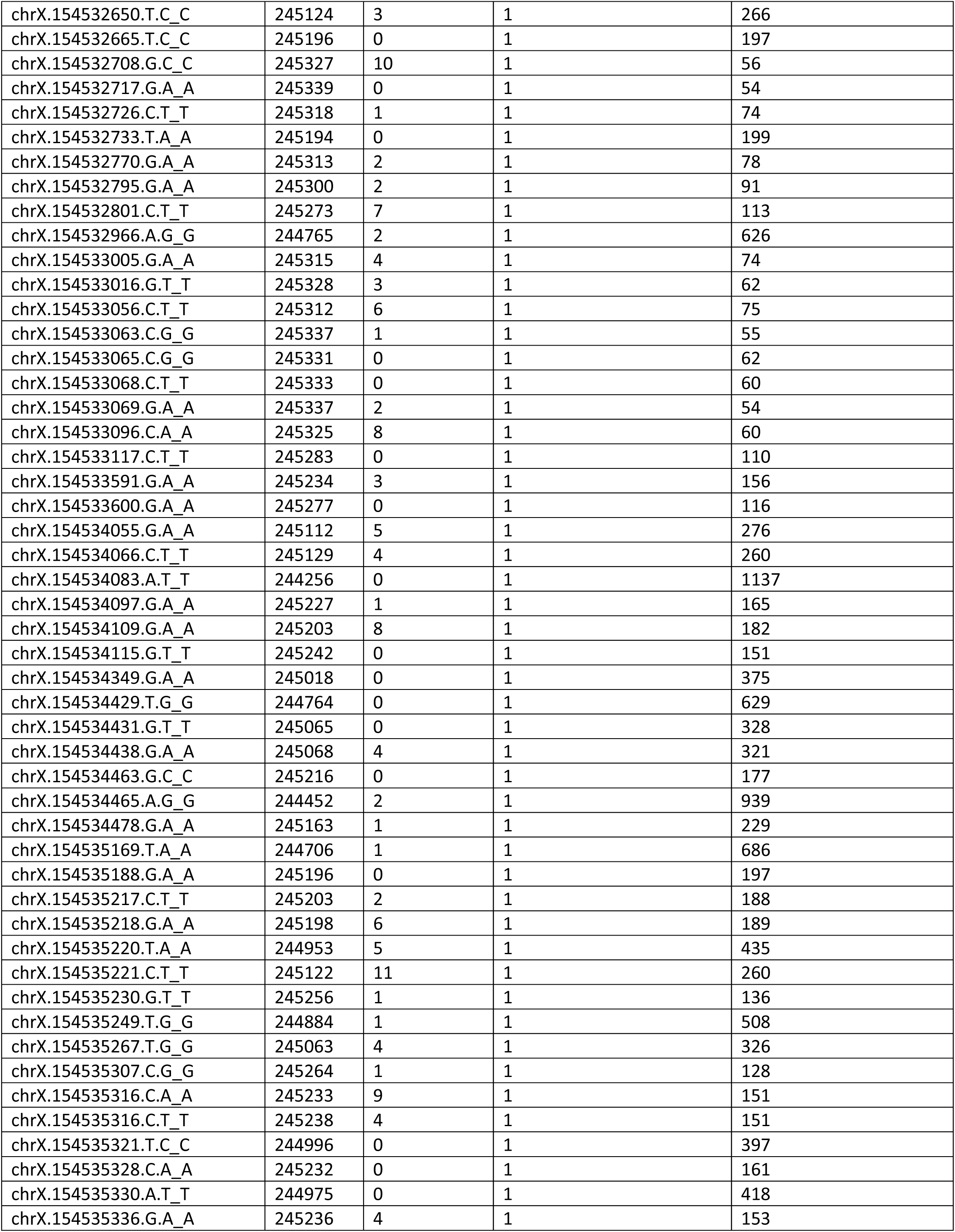
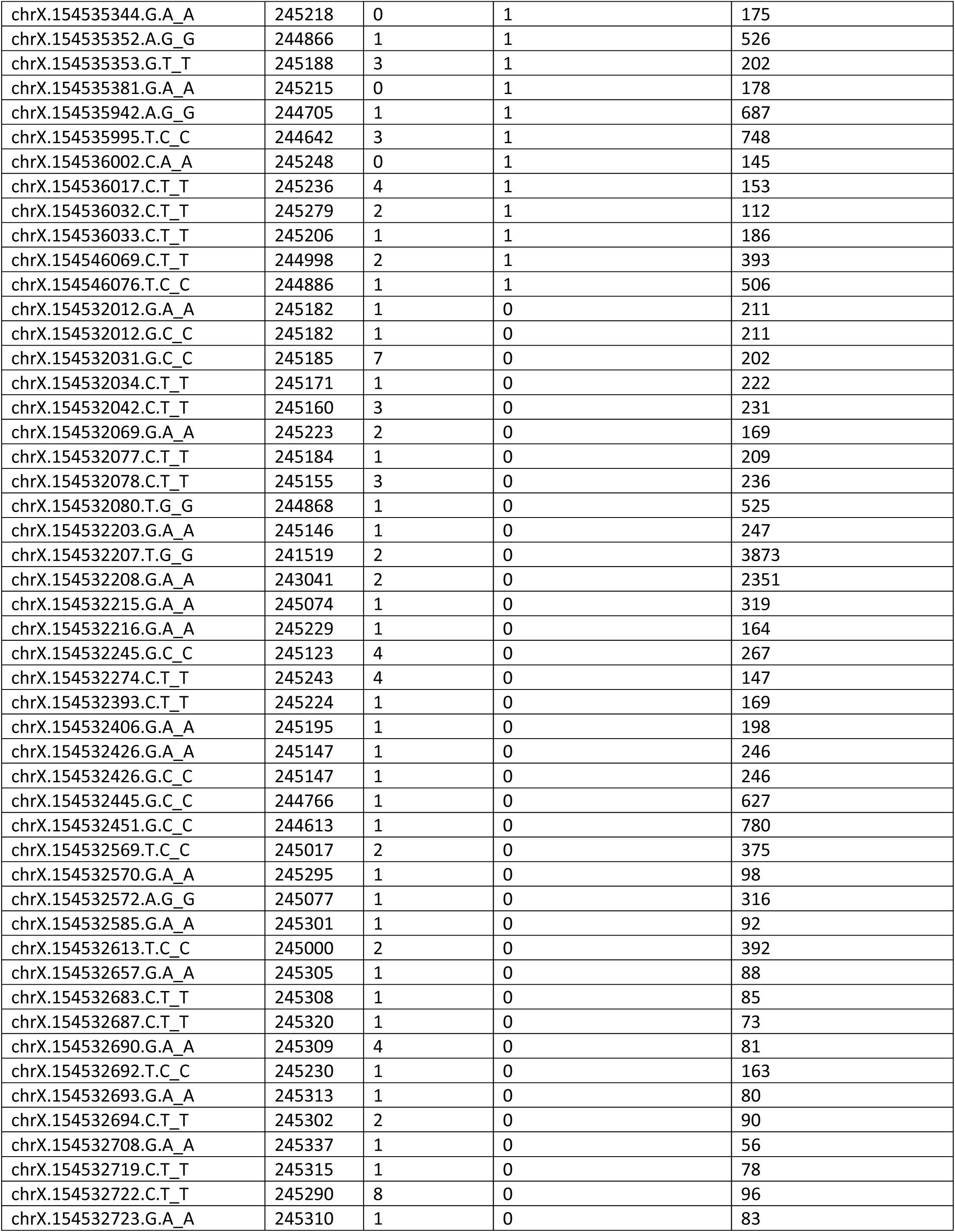
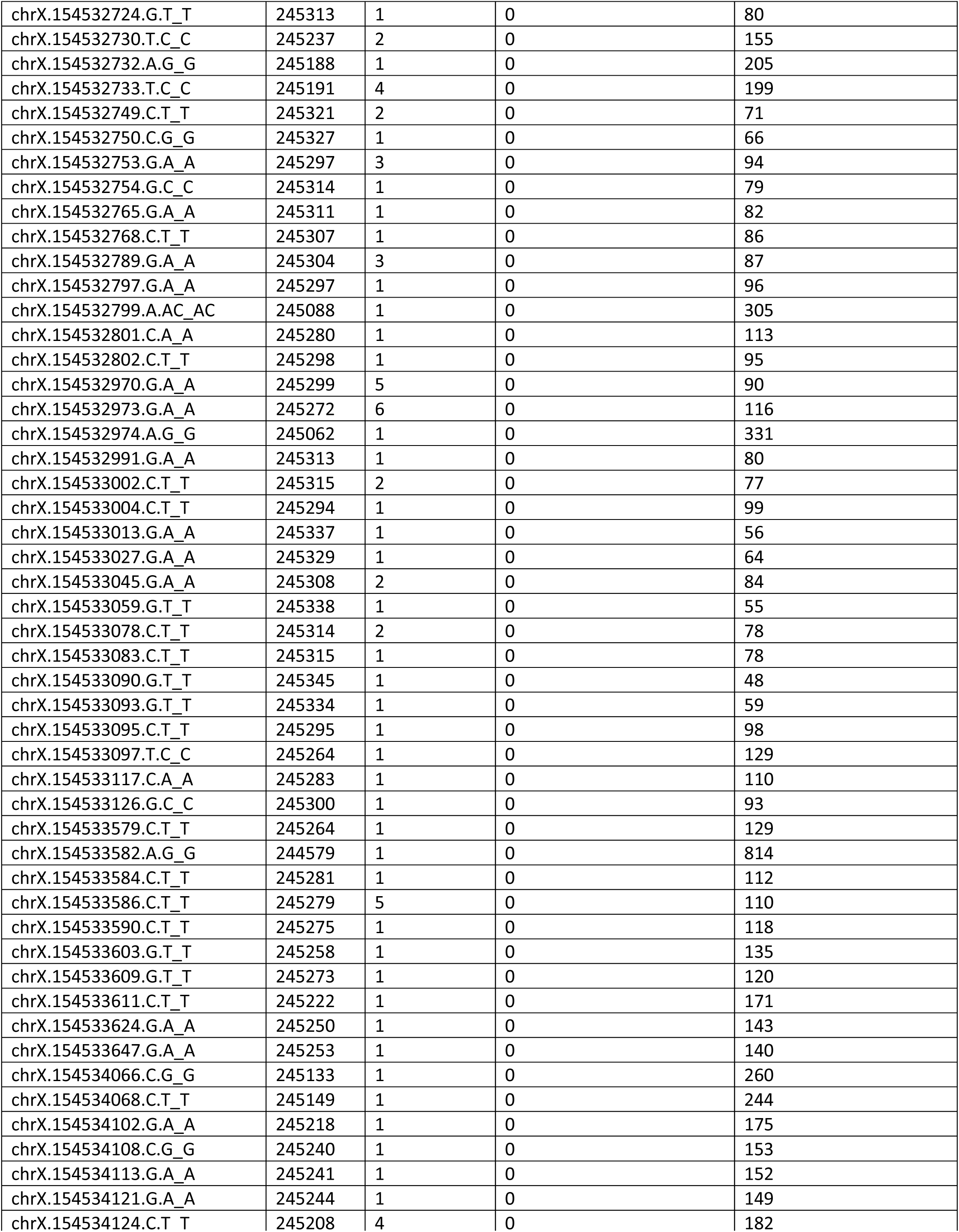
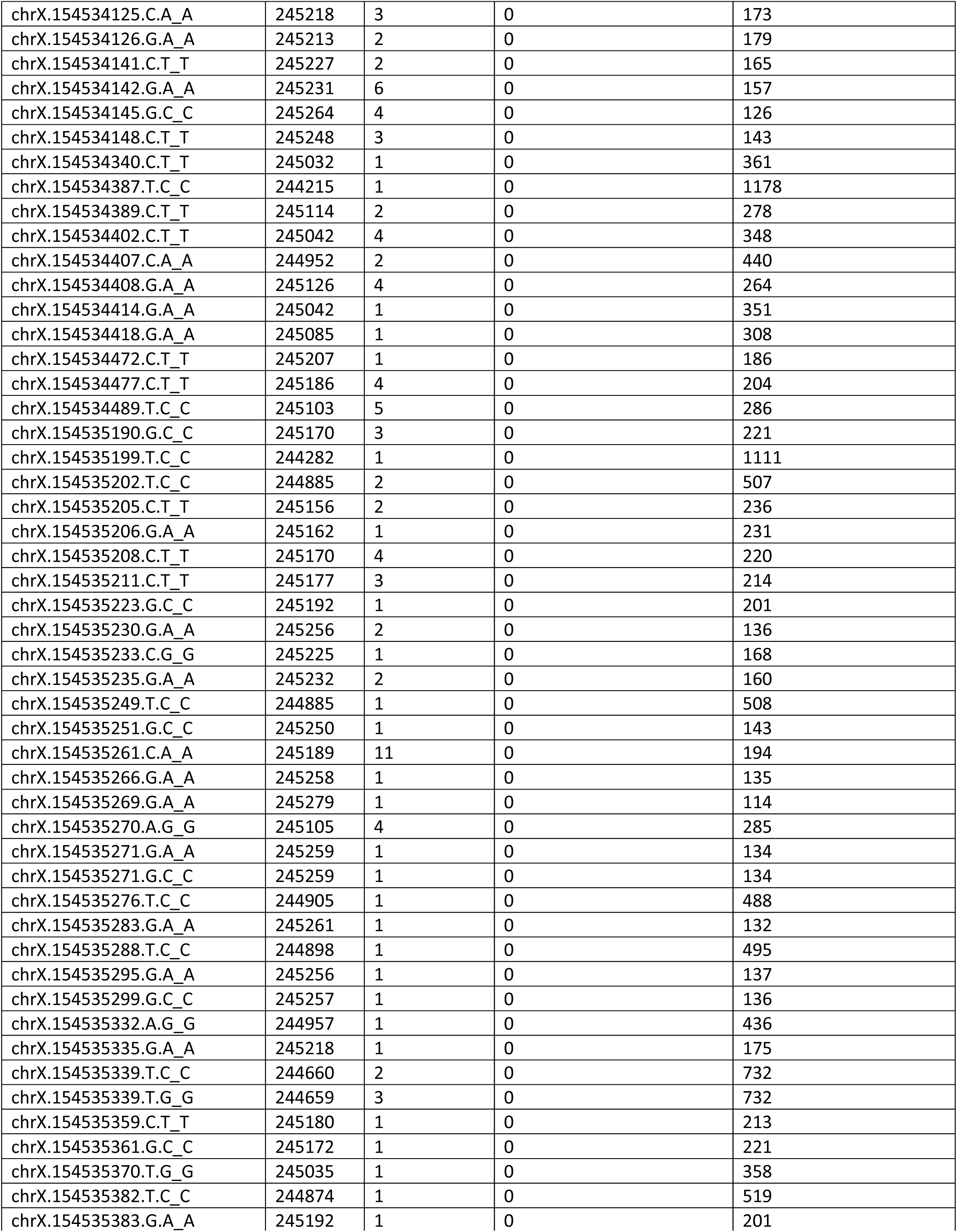
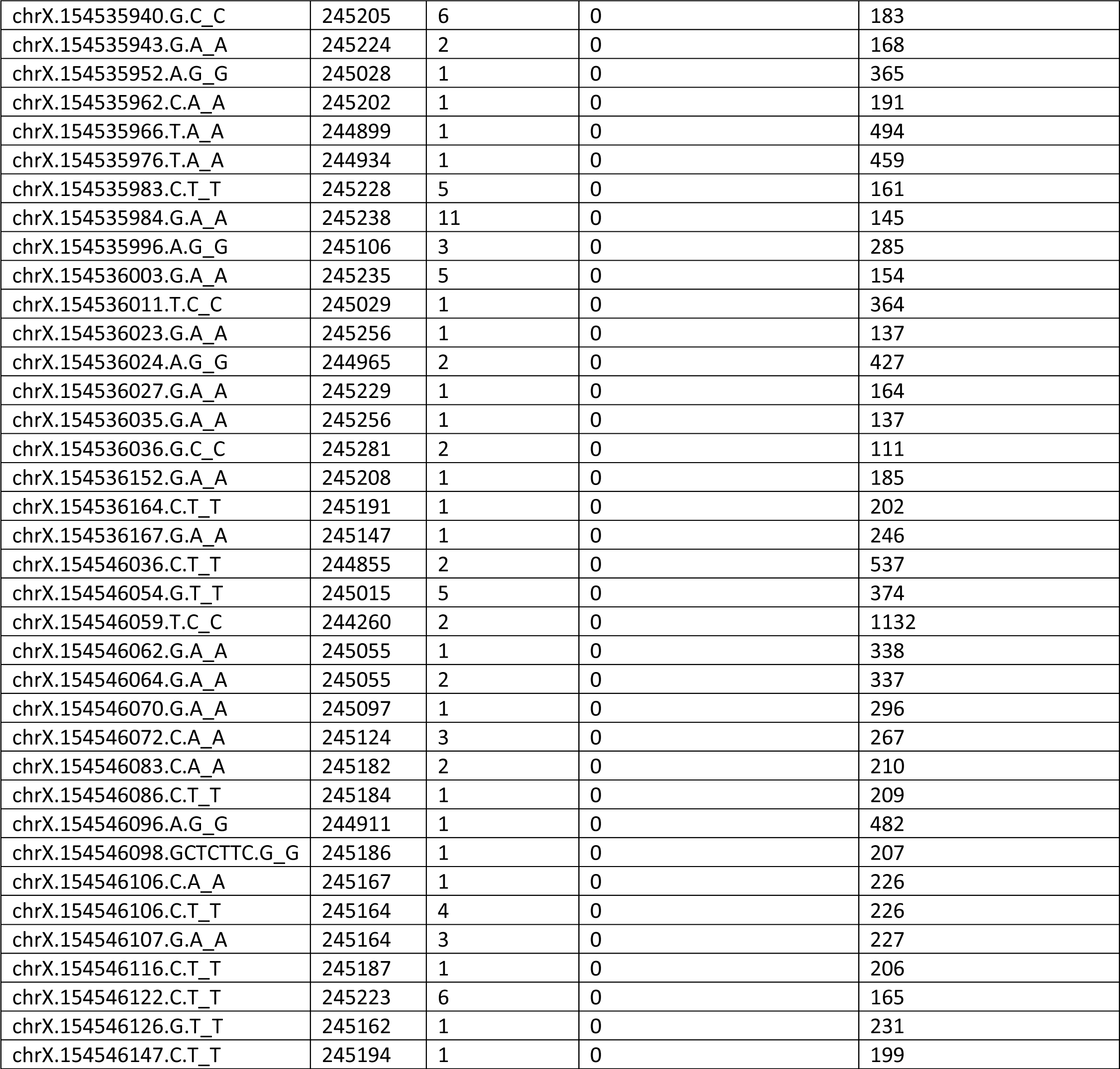
All 359 variants identified in the All of Us Research Project participants in the G6PD CDS. *The nucleotide listed after the “_” is the counted minor allele.

**Supplemental Table S7:** Nonsynonymous SNPs found in the All of Us G6PD CDS that were not previously annotated by Geck. et al. Computational predictors (“Extra”) are default outputs of the UCSC variant annotation integrator tool for multiple transcripts.^23,34^

| snp | Consequence | Position. in.CDS | Position.in .protein | Amino.acid .change | Codon.c hange | Co.located. Variation | Extra |
| --- | --- | --- | --- | --- | --- | --- | --- |
| chrX:154532034 :C:T | missense | 1514 | 505 | G/D | gGc/gAc | - | SIFT=T(0.301,,0.176,0.251);PP2HVAR=P(0.801);PP2HDIV=D(0.989);EXON=13/13 |
| chrX:154532077 :C:T | missense | 1471 | 491 | E/K | Gag/Aag | - | SIFT=T(0.058,,0.056,0.037);EXON=13/13 |
| chrX:154532207 :T:G | missense | 1438 | 480 | I/L | Atc/Ctc | - | SIFT=T(0.469,,0.552,0.444);PP2HVAR=B(0.007);PP2HDIV=B(0.0);EXON=12/13 |
| chrX:154532216 :G:A | missense | 1429 | 477 | P/S | Ccc/Tcc | - | SIFT=T(0.611,,0.821,0.867);PP2HVAR=B(0.003);PP2HDIV=B(0.0);EXON=12/13 |
| chrX:154532393<br>:C:T | missense | 1357 | 453 | V/M | Gtg/Atg | - | SIFT=D(0.0,,0.0,0.0);PP2HVAR=D(0.994);PP2HDIV=D(1.0);<br>EXON=11/13 |
| chrX:154532572<br>:A:G | missense | 1282 | 428 | Y/H | Tac/Cac | - | SIFT=D(0.0,,0.0,0.0);PP2HVAR=D(0.994);PP2HDIV=D(1.0);<br>EXON=10/13 |
| chrX:154532719<br>:C:T | missense | 1135 | 379 | D/N | Gac/Aac | - | SIFT=T(0.213,,0.248,0.173);PP2HVAR=B(0.155);PP2HDIV=<br>B(0.273);EXON=10/13 |
| chrX:154532724<br>:G:T | missense | 1130 | 377 | A/D | gCc/gAc | - | SIFT=T(0.064,,0.031,0.077);PP2HVAR=B(0.013);PP2HDIV=<br>B(0.006);EXON=10/13 |
| chrX:154532754<br>:G:C | missense | 1100 | 367 | A/G | gCc/gGc | - | SIFT=D(0.023,,0.025,0.022);PP2HVAR=B(0.295);PP2HDIV<br>=B(0.176);EXON=10/13 |
| chrX:154533013<br>:G:A | missense | 980 | 327 | T/M | aCg/aTg | - | SIFT=D(0.0,,0.0,0.0,0.0);PP2HVAR=D(0.978);PP2HDIV=D(<br>1.0);EXON=9/13 |
| chrX:154533059<br>:G:T | missense | 934 | 312 | P/T | Ccc/Acc | - | SIFT=T(0.09,,0.1,0.072,0.071,0.07);PP2HVAR=B(0.231);PP<br>2HDIV=B(0.035);EXON=9/13 |
| chrX:154533083<br>:C:T | missense | 910 | 304 | V/I | Gtc/Atc | - | SIFT=T(0.211,,0.241,0.196,0.233,0.25);PP2HVAR=B(0.244<br>);PP2HDIV=B(0.157);EXON=9/13 |
| chrX:154533088<br>:T:C | missense | 905 | 302 | N/S | aAt/aGt | - | SIFT=D(0.004,,0.004,0.004,0.004,0.004);PP2HVAR=B(0.10<br>9);PP2HDIV=B(0.026);EXON=9/13 |
| chrX:154533097<br>:T:C | missense | 896 | 299 | Q/R | cAg/cGg | - | SIFT=T(0.291,,0.296,0.287,0.175,0.147);PP2HVAR=B(0.00<br>1);PP2HDIV=B(0.0);EXON=9/13 |
| chrX:154533117<br>:C:A | missense | 876 | 292 | L/F | ttG/ttT | - | SIFT=D(0.001,,0.001,0.001,0.001,0.001);PP2HVAR=D(0.99<br>9);PP2HDIV=D(1.0);EXON=9/13 |
| chrX:154533584<br>:C:T | missense | 856 | 286 | D/N | Gat/Aat | - | SIFT=T(0.195,,0.197,0.173,0.193,0.175);PP2HVAR=B(0.03<br>3);PP2HDIV=B(0.006);EXON=8/13 |
| chrX:154534083<br>:A:T | missense | 722 | 241 | F/Y | tTt/tAt | - | SIFT=D(0.0,,0.0,0.0,0.003,0.003,0.005);PP2HVAR=D(0.923<br>);PP2HDIV=P(0.946);EXON=7/13 |
| chrX:154534113<br>:G:A | missense | 692 | 231 | A/V | gCc/gTc | - | SIFT=T(0.055,,0.063,0.038,0.047,0.049,0.007);PP2HVAR=<br>B(0.13);PP2HDIV=B(0.27);EXON=7/13 |
| chrX:154534431<br>:G:T | missense | 551 | 184 | S/Y | tCc/tAc | rs78231557<br>2 | SIFT=D(0.001,,0.001,0.001,0.001,0.001,0.0);PP2HVAR=D(<br>0.996);PP2HDIV=D(1.0);EXON=6/13 |
| chrX:154535199<br>:T:C | missense | 454 | 152 | K/E | Aag/Gag | - | SIFT=T(0.362,,0.402,0.345,0.393,0.393,0.632);EXON=5/1<br>3 |
| chrX:154535223<br>:G:C | missense | 430 | 144 | P/A | Ccg/Gcg | - | See Supplemental Table S12 |
| chrX:154535271<br>:G:C | missense | 382 | 128 | L/V | Ctc/Gtc | - | SIFT=T(0.193,,0.176,0.148,0.131,0.129,0.083);PP2HVAR=<br>B(0.144);PP2HDIV=P(0.502);EXON=5/13 |
| chrX:154535276<br>:T:C | missense | 377 | 126 | N/S | aAt/aGt | - | SIFT=T(0.574,,0.478,0.549,0.562,0.573,1.0);PP2HVAR=B(<br>0.0);PP2HDIV=B(0.0);EXON=5/13 |
| chrX:154535299<br>:G:C | stop_gain<br>ed | 354 | 118 | Y/* | taC/taG | - | EXON=5/13 |
| chrX:154535307<br>:C:G | missense | 346 | 116 | A/P | Gcc/Ccc | - | SIFT=T(0.062,,0.052,0.064,0.077,0.084,0.23);PP2HVAR=P<br>(0.452);PP2HDIV=P(0.91);EXON=5/13 |
| chrX:154535381<br>:G:A | missense | 272 | 91 | T/I | aCc/aTc | - | SIFT=T(0.116,,0.12,0.082,0.106,0.059,0.06);PP2HVAR=B(<br>0.024);PP2HDIV=B(0.022);EXON=5/13 |
| chrX:154535942<br>:A:G | missense | 262 | 88 | F/L | Ttc/Ctc | - | SIFT=T(1.0,,1.0,1.0,1.0,1.0,0.664);PP2HVAR=B(0.004);PP2<br>HDIV=B(0.0);EXON=4/13 |
| chrX:154535962<br>:C:A | missense | 242 | 81 | R/L | cGc/cTc | rs78230826<br>6 | SIFT=T(0.153,,0.151,0.149,0.143,0.14,0.088);PP2HVAR=B<br>(0.019);PP2HDIV=B(0.01);EXON=4/13 |
| chrX:154535966<br>:T:A | missense | 238 | 80 | I/F | Atc/Ttc | - | SIFT=T(0.492,,0.519,0.579,0.42,0.41,0.136);PP2HVAR=B(<br>0.401);PP2HDIV=P(0.609);EXON=4/13 |
| chrX:154546070<br>:G:A | missense | 86 | 29 | S/L | tCg/tTg | - | SIFT=T(0.112,,0.153,0.101,0.101,0.109,0.109);PP2HVAR=<br>B(0.001);PP2HDIV=B(0.001);EXON=2/13 |
| chrX:154546086<br>:C:T | missense | 70 | 24 | D/N | Gat/Aat | - | SIFT=T(0.25,,0.215,0.31,0.227,0.231,0.305);PP2HVAR=B(<br>0.001);PP2HDIV=B(0.0);EXON=2/13 |
| chrX:154546098<br>:GCTCTTC:G | inframe_d<br>eletion | 53-58 | 18 | EEL/V | gAAGAG<br>Ctt/gtt | - | EXON=2/13 |

**Supplemental Table S8:** All variants found in the G6PD gene, including introns, in the 82 individuals with a diagnosis of G6PD deficiency. Note: the A+ reference allele (chrX:154535277:T:C) was counted due to it being the minor allele in this group of 82 individuals.

| ch<br>r | Name of snp | A1_usually_m<br>inor | A2_usually_<br>major | A1_hom/hemi_c<br>ount | het_co<br>unt | A2_hom/hemi_c<br>ount | missi<br>ng |
| --- | --- | --- | --- | --- | --- | --- | --- |
| 23 | chrX:154533413:G:A | G | A | 28 | 23 | 31 | 0 |
| 23 | chrX:154535277:T:C | T | C | 28 | 23 | 31 | 0 |
| 23 | chrX:154536313:T:C | T | C | 28 | 23 | 31 | 0 |
| 23 | chrX:154538285:A:G | A | G | 28 | 18 | 36 | 0 |
| 23 | chrX:154542910:A:G | A | G | 28 | 23 | 31 | 0 |
| 23 | chrX:154547570:T:C | T | C | 28 | 18 | 36 | 0 |
| 23 | chrX:154537970:A:G | A | G | 26 | 18 | 36 | 2 |
| 23 | chrX:154533860:G:A | A | G | 24 | 26 | 32 | 0 |
| 23 | chrX:154536448:C:T | T | C | 24 | 26 | 32 | 0 |
| 23 | chrX:154536519:G:A | A | G | 24 | 26 | 32 | 0 |
| 23 | chrX:154534556:G:C | C | G | 23 | 26 | 33 | 0 |
| 23 | chrX:154536002:C:T | T | C | 23 | 27 | 32 | 0 |
| 23 | chrX:154537990:GT:G | GT | G | 20 | 23 | 33 | 6 |
| 23 | chrX:154531643:C:T | T | C | 14 | 11 | 57 | 0 |
| 23 | chrX:154532293:G:A | A | G | 14 | 11 | 57 | 0 |
| 23 | chrX:154540638:G:GA | GA | G | 14 | 11 | 20 | 37 |
| 23 | chrX:154543081:T:C | C | T | 14 | 11 | 57 | 0 |
| 23 | chrX:154532439:A:G | A | G | 10 | 7 | 65 | 0 |
| 23 | chrX:154543136:A:C | A | C | 8 | 4 | 70 | 0 |
| 23 | chrX:154534419:G:A | A | G | 7 | 3 | 72 | 0 |
| 23 | chrX:154543369:G:A | A | G | 7 | 3 | 72 | 0 |
| 23 | chrX:154538857:G:C | G | C | 6 | 4 | 70 | 2 |
| 23 | chrX:154536951:G:A | A | G | 2 | 2 | 78 | 0 |
| 23 | chrX:154538766:A:G | G | A | 2 | 3 | 77 | 0 |
| 23 | chrX:154532257:C:T | T | C | 1 | 0 | 81 | 0 |
| 23 | chrX:154532390:G:A | A | G | 1 | 0 | 81 | 0 |
| 23 | chrX:154533025:A:G | G | A | 1 | 1 | 80 | 0 |
| 23 | chrX:154534291:C:T | T | C | 1 | 0 | 80 | 1 |
| 23 | chrX:154536782:A:G | G | A | 1 | 4 | 77 | 0 |
| 23 | chrX:154537557:G:C | C | G | 1 | 5 | 76 | 0 |
| 23 | chrX:154531869:G:A | A | G | 0 | 1 | 81 | 0 |
| 23 | chrX:154531928:GTCC:<br>G | G | GTCC | 0 | 5 | 74 | 3 |
| 23 | chrX:154531974:C:T | T | C | 0 | 1 | 0 | 81 |
| 23 | chrX:154532214:G:A | A | G | 0 | 5 | 76 | 1 |
| 23 | chrX:154532738:C:T | T | C | 0 | 5 | 77 | 0 |
| 23 | chrX:154533349:G:A | A | G | 0 | 5 | 77 | 0 |
| 23 | chrX:154534177:G:A | A | G | 0 | 1 | 80 | 1 |
| 23 | chrX:154534387:T:C | C | T | 0 | 1 | 81 | 0 |
| 23 | chrX:154534440:T:A | A | T | 0 | 1 | 80 | 1 |
| 23 | chrX:154534699:G:A | A | G | 0 | 4 | 78 | 0 |
| 23 | chrX:154534853:T:G | G | T | 0 | 1 | 81 | 0 |
| 23 | chrX:154535681:GCT:G | G | GCT | 0 | 1 | 81 | 0 |
| 23 | chrX:154536618:T:C | C | T | 0 | 1 | 78 | 3 |
| 23 | chrX:154537304:TCAA<br>AAAA:T | T | TCAAAAAA | 0 | 1 | 81 | 0 |
| 23 | chrX:154537547:G:A | A | G | 0 | 5 | 77 | 0 |
| 23 | chrX:154537568:T:TTG | TTG | T | 0 | 1 | 81 | 0 |
| 23 | chrX:154537570:AAG:A | A | AAG | 0 | 1 | 79 | 2 |
| 23 | chrX:154537576:C:T | T | C | 0 | 1 | 80 | 1 |
| 23 | chrX:154537582:T:C | C | T | 0 | 1 | 78 | 3 |
| 23 | chrX:154537594:T:G | G | T | 0 | 1 | 81 | 0 |
| 23 | chrX:154537602:C:T | T | C | 0 | 2 | 78 | 2 |
| 23 | chrX:154537990:G:GT | GT | G | 0 | 1 | 75 | 6 |
| 23 | chrX:154537990:GTT:G | G | GTT | 0 | 2 | 74 | 6 |
| 23 | chrX:154539216:G:A | A | G | 0 | 1 | 81 | 0 |
| 23 | chrX:154539241:G:C | C | G | 0 | 1 | 81 | 0 |
| 23 | chrX:154539762:G:A | A | G | 0 | 1 | 81 | 0 |
| 23 | chrX:154539884:G:T | T | G | 0 | 5 | 74 | 3 |
| 23 | chrX:154540235:T:C | C | T | 0 | 1 | 79 | 2 |
| 23 | chrX:154540668:C:T | T | C | 0 | 1 | 70 | 11 |
| 23 | chrX:154541674:G:A | A | G | 0 | 1 | 81 | 0 |
| 23 | chrX:154541831:G:A | A | G | 0 | 1 | 81 | 0 |
| 23 | chrX:154541868:AT:A | A | AT | 0 | 1 | 81 | 0 |
| 23 | chrX:154542980:T:C | C | T | 0 | 5 | 77 | 0 |
| 23 | chrX:154543105:C:T | T | C | 0 | 1 | 81 | 0 |
| 23 | chrX:154543179:G:C | C | G | 0 | 1 | 81 | 0 |
| 23 | chrX:154543869:A:AT | AT | A | 0 | 1 | 72 | 9 |
| 23 | chrX:154544533:G:A | A | G | 0 | 4 | 78 | 0 |
| 23 | chrX:154544629:T:TGG | TGG | T | 0 | 1 | 81 | 0 |
| 23 | chrX:154544847:C:T | T | C | 0 | 5 | 77 | 0 |
| 23 | chrX:154545836:C:CA | CA | C | 0 | 5 | 53 | 24 |

**Supplemental Table S9:** Fisher exact test results comparing the allele frequencies of all *G6PD* variants (chrX:154531391-154547572) found in 19 individuals with a deficiency diagnosis who didn’t have WHO Class I/II/III or A/B variants (excluding c.1311C>T) versus 1097 individuals who had normal (> 60%) G6PD activity and no WHO class I/II/III or class A/B variant (excluding c.1311C>T). *The Bonferroni significance threshold is 0.0007 (0.05/70), thus no variants were statistically significant in this analysis.

| SNP | A1 | A1 Case Freq | A1 Control Freq | A2 | Fisher P* | OR |
| --- | --- | --- | --- | --- | --- | --- |
| chrX:154536951:G:A | A | 0.1562 | 0.02958 | G | 0.002871 | 6.075 |
| chrX:154538766:A:G | G | 0.1562 | 0.04707 | A | 0.01785 | 3.749 |
| chrX:154544629:T:TGG | TGG | 0.03125 | 0 | T | 0.01824 | NA |
| chrX:154534387:T:C | C | 0.03125 | 0 | T | 0.01826 | NA |
| chrX:154539762:G:A | A | 0.03125 | 0 | G | 0.01829 | NA |
| chrX:154536782:A:G | G | 0.125 | 0.04128 | A | 0.04489 | 3.318 |
| chrX:154536618:T:C | C | 0.03125 | 0.004684 | T | 0.1542 | 6.855 |
| chrX:154538857:G:C | G | 0.03125 | 0.118 | C | 0.1668 | 0.2411 |
| chrX:154543136:A:C | A | 0.03125 | 0.1201 | C | 0.1668 | 0.2364 |
| chrX:154531869:G:A | A | 0.03125 | 0.006384 | G | 0.1987 | 5.021 |
| chrX:154539216:G:A | A | 0.03125 | 0.006388 | G | 0.1988 | 5.018 |
| chrX:154531643:C:T | C | 0.2812 | 0.3757 | T | 0.357 | 0.6502 |
| chrX:154532293:G:A | G | 0.2812 | 0.3761 | A | 0.357 | 0.6492 |
| chrX:154543081:T:C | T | 0.2812 | 0.3748 | C | 0.357 | 0.6528 |
| chrX:154531928:GTCC:G | G | 0 | 0.0457 | GTCC | 0.3975 | 0 |
| chrX:154532738:C:T | T | 0 | 0.04466 | C | 0.3975 | 0 |
| chrX:154533349:G:A | A | 0 | 0.04466 | G | 0.3975 | 0 |
| chrX:154538285:A:G | G | 0.0625 | 0.1236 | A | 0.4179 | 0.4726 |
| chrX:154547570:T:C | C | 0.0625 | 0.1237 | T | 0.4179 | 0.4723 |
| chrX:154537970:A:G | G | 0.0625 | 0.1286 | A | 0.4194 | 0.4516 |
| chrX:154539241:G:C | C | 0.03125 | 0.01801 | G | 0.4484 | 1.759 |
| chrX:154537602:C:T | T | 0.03226 | 0.02214 | C | 0.5064 | 1.472 |
| chrX:154537557:G:C | C | 0.03125 | 0.0244 | G | 0.5515 | 1.29 |
| chrX:154544847:C:T | T | 0 | 0.04002 | C | 0.6346 | 0 |
| chrX:154542980:T:C | C | 0 | 0.04067 | T | 0.6363 | 0 |
| chrX:154539884:G:T | T | 0 | 0.04179 | G | 0.6394 | 0 |
| chrX:154537990:GT:G | G | 0.1379 | 0.1792 | GT | 0.8061 | 0.7328 |
| chrX:154531974:C:T | T | NA | 0.3333 | C | 1 | NA |
| chrX:154532214:G:A | A | 0 | 0.03027 | G | 1 | 0 |
| chrX:154532257:C:T | T | 0 | NA | C | 1 | NA |
| chrX:154532390:G:A | A | 0 | NA | G | 1 | NA |
| chrX:154532439:A:G | A | 0.1875 | 0.1897 | G | 1 | 0.9859 |
| chrX:154533025:A:G | G | 0 | 0 | A | 1 | NA |
| chrX:154533413:G:A | A | 0.0625 | 0.06903 | G | 1 | 0.8992 |
| chrX:154533860:G:A | A | 0 | 0.01334 | G | 1 | 0 |
| chrX:154534177:G:A | A | 0 | 0.01044 | G | 1 | 0 |
| chrX:154534291:C:T | T | 0 | NA | C | 1 | NA |
| chrX:154534419:G:A | A | 0 | 0 | G | 1 | NA |
| chrX:154534440:T:A | A | 0 | 0 | T | 1 | NA |
| chrX:154534556:G:C | C | 0 | 0.01336 | G | 1 | 0 |
| chrX:154534699:G:A | A | 0 | 0.01856 | G | 1 | 0 |
| chrX:154534853:T:G | G | 0 | 0 | T | 1 | NA |
| chrX:154535277:T:C | C | 0.0625 | 0.07023 | T | 1 | 0.8826 |
| chrX:154535681:GCT:G | G | 0 | 0.0058 | GCT | 1 | 0 |
| chrX:154536002:C:T | T | 0 | 0 | C | 1 | NA |
| chrX:154536313:T:C | C | 0.0625 | 0.07599 | T | 1 | 0.8107 |
| chrX:154536448:C:T | T | 0 | 0.01336 | C | 1 | 0 |
| chrX:154536519:G:A | A | 0 | 0.01337 | G | 1 | 0 |
| chrX:154537304:TCAAAAAA:T | T | 0 | NA | TCAAAAAA | 1 | NA |
| chrX:154537547:G:A | A | 0 | 0.02695 | G | 1 | 0 |
| chrX:154537568:T:TTG | TTG | 0 | NA | T | 1 | NA |
| chrX:154537570:AAG:A | A | 0 | NA | AAG | 1 | NA |
| chrX:154537576:C:T | T | 0 | NA | C | 1 | NA |
| chrX:154537582:T:C | C | 0 | NA | T | 1 | NA |
| chrX:154537594:T:G | G | 0 | NA | T | 1 | NA |
| chrX:154537990:G:GT | GT | 0 | 0.02462 | G | 1 | 0 |
| chrX:154537990:GTT:G | G | 0 | 0.000683 | GTT | 1 | 0 |
| chrX:154540235:T:C | C | 0 | 0 | T | 1 | NA |
| chrX:154540638:G:GA | G | 0.2069 | 0.219 | GA | 1 | 0.9303 |
| chrX:154540668:C:T | T | 0 | 0.001367 | C | 1 | 0 |
| chrX:154541674:G:A | A | 0 | 0.005804 | G | 1 | 0 |
| chrX:154541831:G:A | A | 0 | 0.001743 | G | 1 | 0 |
| chrX:154541868:AT:A | A | 0 | 0.008716 | AT | 1 | 0 |
| chrX:154542910:A:G | G | 0.0625 | 0.07367 | A | 1 | 0.8383 |
| chrX:154543105:C:T | T | 0 | 0.01914 | C | 1 | 0 |
| chrX:154543179:G:C | C | 0 | NA | G | 1 | NA |
| chrX:154543369:G:A | A | 0 | 0 | G | 1 | NA |
| chrX:154543869:A:AT | AT | 0 | 0.0165 | A | 1 | 0 |
| chrX:154544533:G:A | A | 0 | 0.02382 | G | 1 | 0 |
| chrX:154545836:C:CA | CA | 0.04 | 0.05763 | C | 1 | 0.6814 |

**Supplemental Table S10:** Linear regression results for 58 intron and UTR variants present in 2 or fewer alleles and carried by at least 1 homozygote or hemizygote. No variants passed the Bonferroni multiple testing threshold.

| CHR | SNP | BP | A1 | TEST | NMISS | BETA | STAT | P |
| --- | --- | --- | --- | --- | --- | --- | --- | --- |
| 23 | chrX:154539082:T:C | 154539082 | C | ADD | 1245 | -0.4041 | -2.035 | 0.04205 |
| 23 | chrX:154537101:C:T | 154537101 | T | ADD | 1229 | -0.4242 | -1.905 | 0.05697 |
| 23 | chrX:154538874:C:T | 154538874 | T | ADD | 1230 | -0.3679 | -1.845 | 0.06525 |
| 23 | chrX:154535853:TG:T | 154535853 | T | ADD | 1236 | 0.3974 | 1.793 | 0.07329 |
| 23 | chrX:154531710:C:T | 154531710 | T | ADD | 1233 | 0.3977 | 1.788 | 0.07402 |
| 23 | chrX:154544161:T:TTTTTG | 154544161 | TTTTTG | ADD | 753 | -0.3661 | -1.641 | 0.1013 |
| 23 | chrX:154541926:G:A | 154541926 | A | ADD | 1248 | 0.3426 | 1.544 | 0.1228 |
| 23 | chrX:154541582:AGC:A | 154541582 | A | ADD | 1244 | -0.3269 | -1.474 | 0.1407 |
| 23 | chrX:154541592:AGAG:A | 154541592 | A | ADD | 1244 | -0.3265 | -1.474 | 0.1407 |
| 23 | chrX:154541589:T:G | 154541589 | G | ADD | 1245 | -0.3268 | -1.474 | 0.1409 |
| 23 | chrX:154541586:T:C | 154541586 | C | ADD | 1246 | -0.3266 | -1.472 | 0.1413 |
| 23 | chrX:154541590:C:A | 154541590 | A | ADD | 1246 | -0.3266 | -1.472 | 0.1413 |
| 23 | chrX:154541596:T:C | 154541596 | C | ADD | 1246 | -0.3265 | -1.471 | 0.1415 |
| 23 | chrX:154538129:G:A | 154538129 | A | ADD | 1242 | -0.3179 | -1.434 | 0.1518 |
| 23 | chrX:154544904:G:A | 154544904 | A | ADD | 1245 | -0.3048 | -1.372 | 0.1702 |
| 23 | chrX:154545153:C:T | 154545153 | T | ADD | 1247 | 0.2959 | 1.333 | 0.1828 |
| 23 | chrX:154533488:T:C | 154533488 | C | ADD | 1244 | -0.2833 | -1.276 | 0.2024 |
| 23 | chrX:154543326:G:T | 154543326 | T | ADD | 1245 | -0.2797 | -1.259 | 0.2081 |
| 23 | chrX:154545928:A:C | 154545928 | C | ADD | 1236 | -0.2767 | -1.246 | 0.2129 |
| 23 | chrX:154540252:C:T | 154540252 | T | ADD | 1228 | 0.2333 | 1.173 | 0.241 |
| 23 | chrX:154546822:AG:A | 154546822 | A | ADD | 1229 | 0.2609 | 1.171 | 0.2416 |
| 23 | chrX:154537546:C:T | 154537546 | T | ADD | 1208 | -0.2557 | -1.144 | 0.253 |
| 23 | chrX:154542355:T:G | 154542355 | G | ADD | 1246 | 0.2369 | 1.067 | 0.2864 |
| 23 | chrX:154546782:C:G | 154546782 | G | ADD | 1247 | -0.1557 | -0.9917 | 0.3215 |
| 23 | chrX:154540668:C:T | 154540668 | T | ADD | 1050 | 0.1542 | 0.9737 | 0.3304 |
| 23 | chrX:154541524:G:A | 154541524 | A | ADD | 1248 | -0.193 | -0.9716 | 0.3314 |
| 23 | chrX:154531974:C:T | 154531974 | T | ADD | 4 | -0.5079 | -1.009 | 0.4191 |
| 23 | chrX:154533208:A:T | 154533208 | T | ADD | 1247 | -0.1779 | -0.801 | 0.4233 |
| 23 | chrX:154541217:C:A | 154541217 | A | ADD | 1247 | 0.1698 | 0.7647 | 0.4446 |
| 23 | chrX:154535088:C:T | 154535088 | T | ADD | 1248 | -0.1668 | -0.7515 | 0.4525 |
| 23 | chrX:154543149:G:GA | 154543149 | GA | ADD | 1247 | -0.1668 | -0.7512 | 0.4526 |
| 23 | chrX:154538166:TG:T | 154538166 | T | ADD | 1242 | -0.1669 | -0.7509 | 0.4529 |
| 23 | chrX:154543720:C:T | 154543720 | T | ADD | 1245 | -0.1408 | -0.7097 | 0.478 |
| 23 | chrX:154532314:G:A | 154532314 | A | ADD | 1248 | 0.1566 | 0.7054 | 0.4807 |
| 23 | chrX:154539029:T:C | 154539029 | C | ADD | 1244 | -0.1562 | -0.7025 | 0.4825 |
| 23 | chrX:154541987:C:G | 154541987 | G | ADD | 1246 | -0.154 | -0.6932 | 0.4883 |
| 23 | chrX:154540123:C:A | 154540123 | A | ADD | 1230 | 0.1476 | 0.664 | 0.5068 |
| 23 | chrX:154536198:T:A | 154536198 | A | ADD | 1244 | -0.1358 | -0.611 | 0.5413 |
| 23 | chrX:154545604:G:A | 154545604 | A | ADD | 1247 | -0.1161 | -0.523 | 0.601 |
| 23 | chrX:154539722:T:C | 154539722 | C | ADD | 1236 | -0.1 | -0.4495 | 0.6531 |
| 23 | chrX:154535414:G:C | 154535414 | C | ADD | 1247 | 0.08016 | 0.4033 | 0.6868 |
| 23 | chrX:154535756:G:A | 154535756 | A | ADD | 1248 | 0.08013 | 0.4033 | 0.6868 |
| 23 | chrX:154536224:C:T | 154536224 | T | ADD | 1248 | -0.08763 | -0.3947 | 0.6932 |
| 23 | chrX:154533144:G:A | 154533144 | A | ADD | 1247 | 0.07553 | 0.34 | 0.7339 |
| 23 | chrX:154535484:G:A | 154535484 | A | ADD | 1246 | 0.07359 | 0.3314 | 0.7404 |
| 23 | chrX:154536543:T:C | 154536543 | C | ADD | 1244 | -0.0504 | -0.3211 | 0.7482 |
| 23 | chrX:154541028:C:T | 154541028 | T | ADD | 1246 | -0.06036 | -0.2719 | 0.7857 |
| 23 | chrX:154543116:C:T | 154543116 | T | ADD | 1246 | 0.04855 | 0.2442 | 0.8071 |
| 23 | chrX:154540180:A:G | 154540180 | G | ADD | 1232 | -0.049 | -0.2211 | 0.825 |
| 23 | chrX:154546628:G:A | 154546628 | A | ADD | 1246 | -0.03668 | -0.1651 | 0.8689 |
| 23 | chrX:154538915:C:T | 154538915 | T | ADD | 1232 | -0.03041 | -0.1371 | 0.891 |
| 23 | chrX:154543381:T:C | 154543381 | C | ADD | 1247 | 0.02901 | 0.1306 | 0.8961 |
| 23 | chrX:154545875:C:CACACCAGG | 154545875 | CACACCAGG | ADD | 1231 | 0.02868 | 0.1289 | 0.8975 |
| 23 | chrX:154545610:G:T | 154545610 | T | ADD | 1248 | -0.02857 | -0.1286 | 0.8977 |
| 23 | chrX:154536124:T:C | 154536124 | C | ADD | 1246 | -0.02507 | -0.1129 | 0.9101 |
| 23 | chrX:154538273:G:A | 154538273 | A | ADD | 1246 | -0.0189 | -0.08504 | 0.9322 |
| 23 | chrX:154539005:T:C | 154539005 | C | ADD | 1239 | -0.01856 | -0.08343 | 0.9335 |
| 23 | chrX:154531975:G:A | 154531975 | A | ADD | 1248 | -0.01627 | -0.07329 | 0.9416 |

**Supplemental Table S11:** Optical density at 600nM of *zwf1Δ S. cerevisiae* expressing *G6PD* alleles Provided as file attachment “Supplemental_Table_S11.txt”.

**Supplemental Table S12:** In silico variant effect predictions for selected variants seen in All of Us, accessed through dbNSFP (last updated March 13, 2024). Scores interpreted according to ClinGen recommendations for PP3/BP4 criteria (PMID 36413997). ENST00000393562 was used for all tools for which it was available (BayesDel, CADD, GERP++, PrimateAI, REVEL, MPC, PolyPhen-2 HumVar), and ENST00000393564 was used for the remaining tools (SIFT, FATHMM, VEST4). Scores from FATHMM and CADD were not included in final analysis because they predicted all tested variants to be pathogenic and benign, respectively. If other tools predicted conflicting evidence for benignity or pathogenicity, consensus was taken when more than ⅔ agreed and no more than one tool dissented, and the consensus was used at supporting level of evidence. If multiple tools provided non-conflicting evidence, the lowest level of support was conservatively used. Provided as file attachment “Supplemental_Table_S12.xlsx”.

